# Plasma p-tau217 predicts cognitive impairments up to ten years before onset in normal older adults

**DOI:** 10.1101/2024.05.09.24307120

**Authors:** Yara Yakoub, Fernando Gonzalez-Ortiz, Nicholas J. Ashton, Christine Déry, Cherie Strikwerda-Brown, Frédéric St-Onge, Valentin Ourry, Michael Schöll, Maiya R. Geddes, Simon Ducharme, Maxime Montembeault, Pedro Rosa-Neto, Jean-Paul Soucy, John C.S. Breitner, Henrik Zetterberg, Kaj Blennow, Judes Poirier, Sylvia Villeneuve, PREVENT-AD Research Group

## Abstract

**Importance:** Positron emission tomography (PET) biomarkers are the gold standard for detection of Alzheimer amyloid and tau *in vivo*. Such imaging can identify cognitively unimpaired (CU) individuals who will subsequently develop cognitive impartment (CI). Plasma biomarkers would be more practical than PET or even cerebrospinal fluid (CSF) assays in clinical settings.

**Objective:** Assess the prognostic accuracy of plasma p-tau217 in comparison to CSF and PET biomarkers for predicting the clinical progression from CU to CI.

**Design:** In a cohort of elderly at high risk of developing Alzheimer’s dementia (AD), we measured the proportion of CU individuals who developed CI, as predicted by Aβ (A+) and/or tau (T+) biomarker assessment from plasma, CSF, and PET. Results from each method were compared with (A-T-) reference individuals. Data were analyzed from June 2023 to April 2024.

**Setting:** Longitudinal observational cohort.

**Participants:** Some 228 participants from the PREVENT-AD cohort were CU at the time of biomarker assessment and had 1 - 10 years of follow-up. Plasma was available from 215 participants, CSF from 159, and amyloid- and tau-PET from 155. Ninety-three participants had assessment using all three methods (main group of interest). Progression to CI was determined by clinical consensus among physicians and neuropsychologists who were blind to plasma, CSF, PET, and MRI findings, as well as *APOE* genotype.

**Exposures:** Plasma Aβ_42/40_ was measured using IP-MS; CSF Aβ _42/40_ using Lumipulse; plasma and CSF p-tau217 using UGOT assay. Aβ-PET employed the ^18^F-NAV4694 ligand, and tau-PET used ^18^F-flortaucipir.

**Main Outcome:** Prognostic accuracy of plasma, CSF, and PET biomarkers for predicting the development of CI in CU individuals.

**Results:** Cox proportional hazard models indicated a greater progression rate in all A+T+ groups compared to A-T-groups (HR = 6.61 [95% CI = 2.06 – 21.17] for plasma, 3.62 [1.49 – 8.81] for CSF and 9.24 [2.34 – 36.43] for PET). The A-T+ groups were small, but also characterized with individuals who developed CI. Plasma biomarkers identified about five times more T+ than PET.

**Conclusion and relevance:** Plasma p-tau217 assessment is a practical method for identification of persons who will develop cognitive impairment up to 10 years later.

**Key Points:** *Question:* Can plasma p-tau217 serve as a prognostic indicator for identifying cognitively unimpaired (CU) individuals at risk of developing cognitive impairments (CI)?

*Findings:* In a longitudinal cohort of CU individuals with a family history of sporadic AD, almost all individuals with abnormal plasma p-tau217 concentrations developed CI within 10 years, regardless of plasma amyloid levels. Similar findings were obtained with CSF p-tau217 and tau-PET. Fluid p-tau217 biomarkers had the main advantage over PET of identifying five times more participants with elevated tau.

*Meaning:* Elevated plasma p-tau217 levels in CU individuals strongly indicate future clinical progression

## Introduction

The protein pathologies that define Alzheimer’s disease (AD) are aggregates of amyloid-beta (Aβ) into plaques, neurofibrillary tangles of hyper-phosphorylated (p) tau, and dystrophic neurites. Aβ and tau start to aggregate years before the onset of the clinical disease. ^1^ The first two protein components can be measured *in vivo* using positron emission tomography (PET), cerebrospinal fluid (CSF) assays and, more recently blood-based biomarkers in plasma. ^2,3^ While PET had recently been found to be valuable at identifying cognitively unimpaired (CU) individuals who developed cognitive impairments (CI) within 3-5 years, ^4,5^ the predictive value of plasma and CSF biomarkers for this purpose is unknown.

Imaging and fluid biomarkers of Aβ and tau reflect different biochemical pools of a given protein; fluid biomarkers capture soluble and diffusible proteins, while PET images capture insoluble aggregates that are characteristic of later stages of the disease.^6^ Consequently, Aβ and tau fluid biomarker abnormalities occur prior to the development of corresponding PET abnormalities,^7–9^ so that positive fluid biomarker results should logically reveal disease pathogenesis earlier before clinical onset than positive PET results.

With the emergence of disease-modifying therapies, it has become urgent to find low-cost and widely available biomarkers that can identify CU who are at imminent risk of developing CI. By the time someone has CI symptoms, the pathology is already well advanced, and Aβ may no longer be the optimal target.^10^ Adverse effects related to anti-amyloid treatments are also more prevalent at more advanced disease stages. ^11^ Prognostic information is therefore of critical value to inform treatment decisions or to establish advance directives.

We assessed the prognostic value of novel plasma and CSF p-tau217 assays as predictors of progression of CU individuals to later CI. We predicted that many CU individuals with abnormal plasma, CSF or PET tau biomarker results would develop CI over a brief interval of years and that plasma and CSF biomarkers would show an advantage over PET by identifying more participants with predictive abnormal tau biomarkers.

## Methods

We included 228 participants from the PREVENT-AD cohort with available plasma (Aβ_42/30_ and p-tau 217), CSF (Aβ_42/40_ and p-tau217) or PET (Aβ and tau PET) biomarkers who were CU when enrolled in the study and at the time of the biomarkers assessment, and who had at least one year of cognitive follow-up upon biomarkers assessment (see Supplement). Overall, 215 participants had plasma, 159 had CSF and 155 had both Aβ- and tau-PET available. For 93 participants, the main group of interest, all biomarkers were available (**Figure 1, eFigure 1** in Supplement). The demographic and clinical information of the 93 participants with all biomarkers can be found in **Table 1**. Demographic characteristics of plasma, CSF and PET full samples can be found in Supplement (**eTable 1**). Written informed consent was obtained from all participants and all research procedures were approved by the Institutional Review Board at McGill University and complied with the ethical principles of the Declaration of Helsinki. A detailed description of the PREVENT-AD cohort is available elsewhere. ^12^

**Figure 1.**
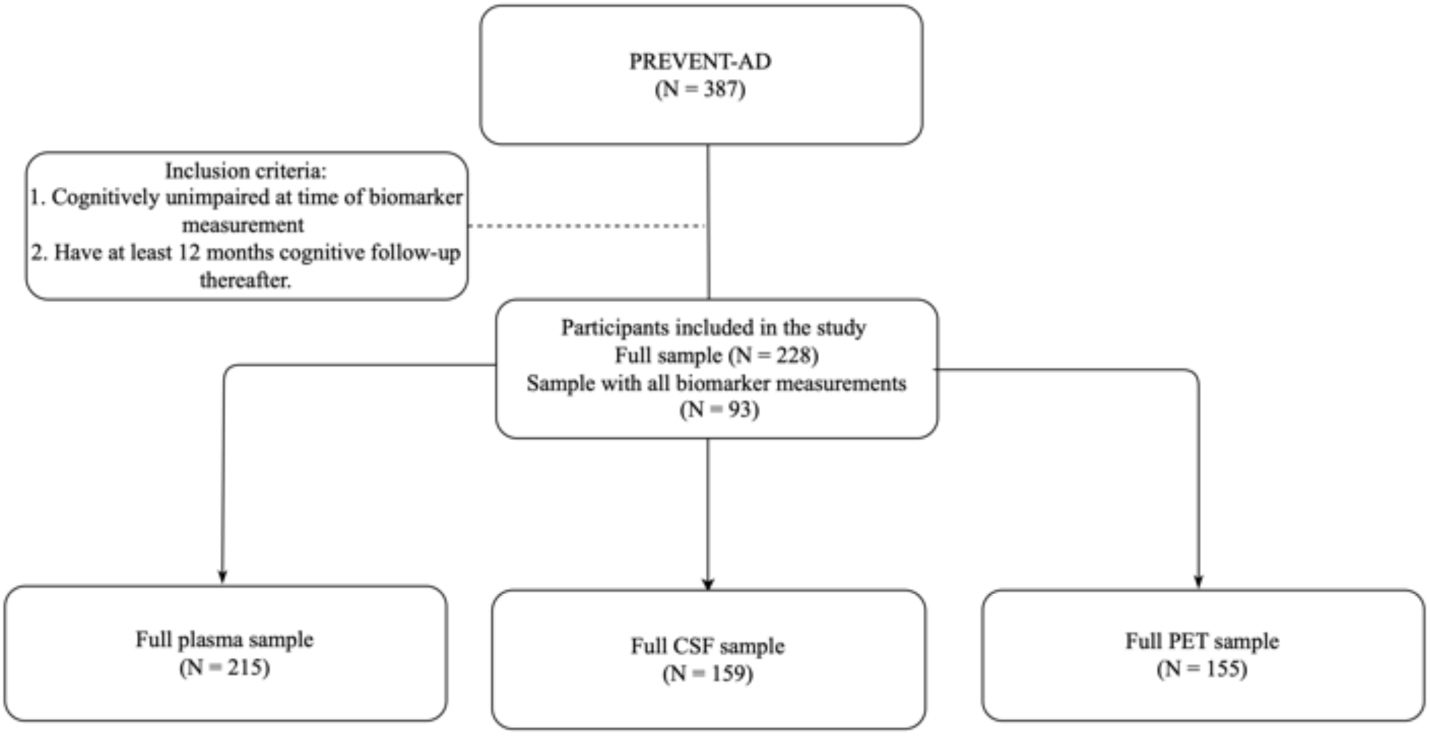
Flowchart of included participants for each plasma, PET and CSF biomarkers sub-samples. We included a total of 215 participants from plasma sample; 159 participants from the CSF sample; and 155 participants from PET sample. A total of 93 participants that have all biomarker measurements were included in the main analyses. Participants were excluded if they did not meet one of the following eligibility criteria: 1). Being cognitively unimpaired at the time of the biomarker measurement, 2). Having at least 12 months of cognitive follow-up thereafter.

**Table 1.**
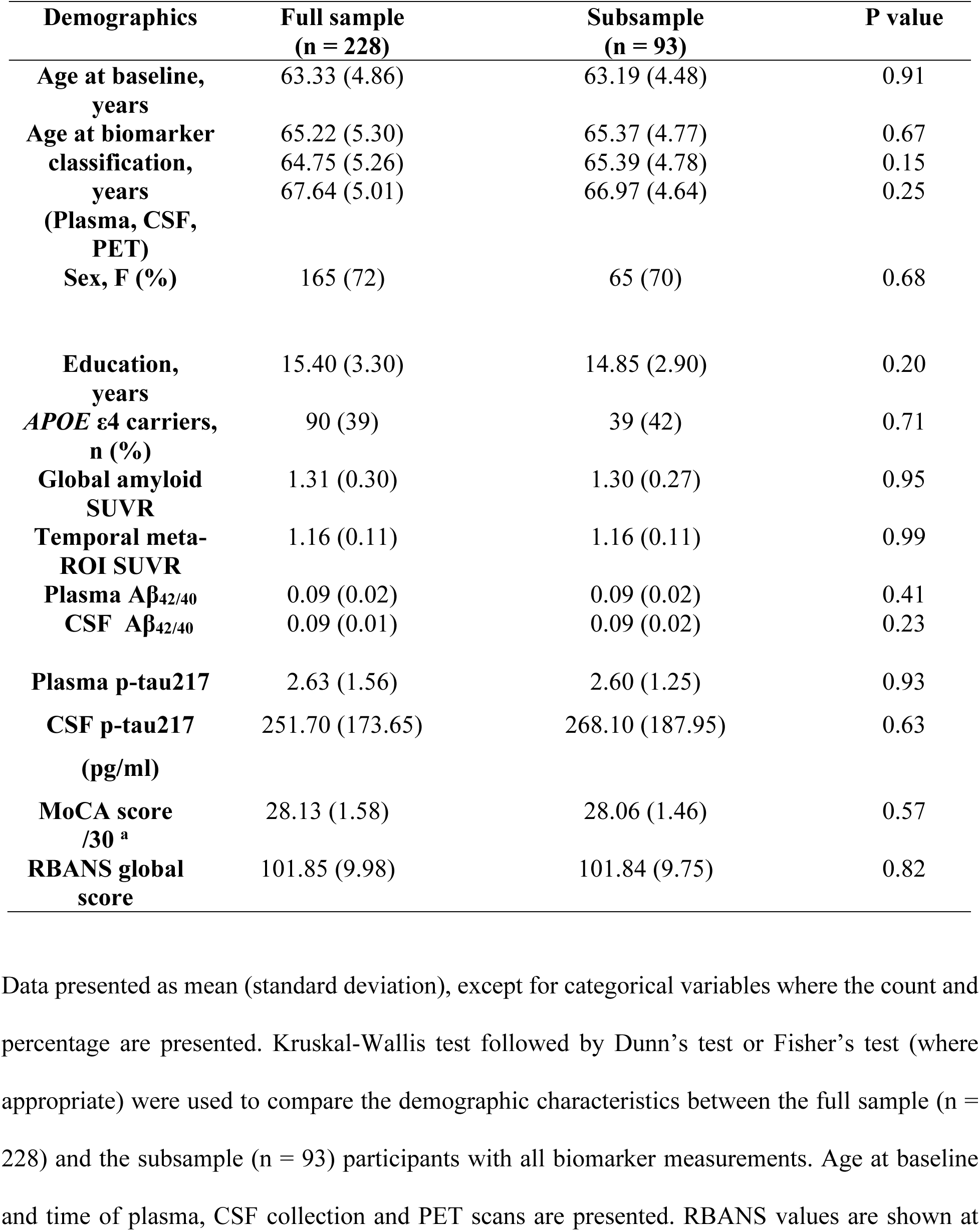
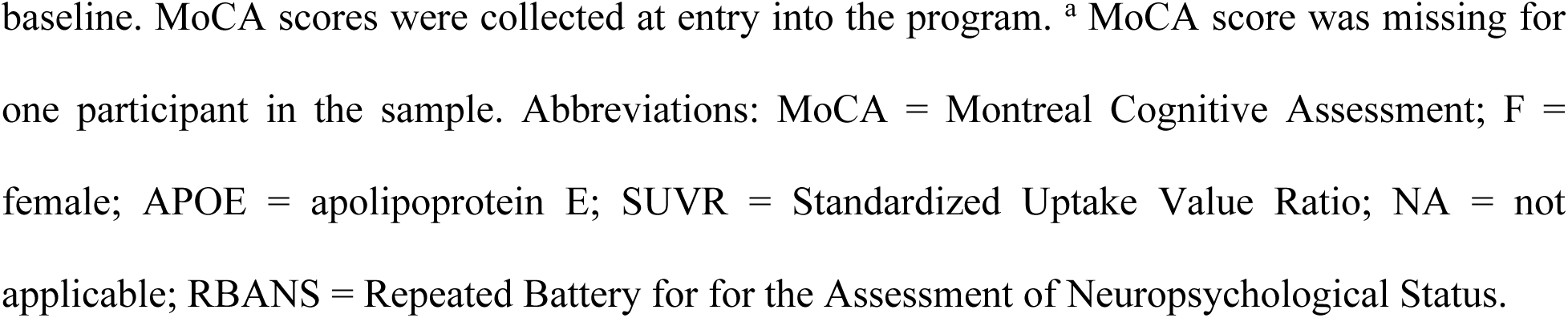
Sample Demographics.

### Clinical outcomes

The cognitive performances of participants whose performance deviated by more than 1 standard deviation from demographically stratified norms on at least one of the five composite subscale score of the Repeatable Battery for the Assessment of Neuropsychological Status (RBANS), on the Trial Making Test, the Stroop, or the Rey-Auditory Verbal Learning Test, were reviewed in multidisciplinary meetings composed off neuropsychologists, neuropsychiatrists, and neurologists. The CI classification was based on an objective decline on all available [between 2 and 10] subjective and objective cognitive evaluations and blind to plasma, CSF, PET, MRI and APOE genotype information. ^4^

### Biomarker measurements and AT classification

Plasma Aβ_40_ and Aβ_42_ concentrations were measured using ultrasensitive immunoprecipitation coupled with mass spectrometry (IP-MS) technique ^13^; CSF Aβ_40_ and Aβ_42_ were analyzed using LUMIPULSE G-automated immunoassay.^14^ Plasma and CSF p-tau217 was measured using an in-house Simoa platform developed at the Clinical Neurochemistry Laboratory, University of Gothenburg.^15^ Aβ -PET was performed using ^18^F-NAV4694 and tau-PET using ^18^F -flortaucipir. ^4^ For the binary analyses Aβ-PET positivity was set at 1.27 SUVR and established based on the global neocortical amyloid-PET retention in the lateral and medial frontal, parietal, and lateral temporal regions of PREVENT-AD participants. Tau-positivity was set at 1.29 SUVR in a temporal meta-ROI, and was established using 2SD from the mean of CU PREVENT-AD Aβ-PET negative individuals.^16^ We used the pre-established thresholds of 0.09 for plasma Aβ_42/40;_ and 0.072 for CSF Aβ_42/40_. ^13,14^ Thresholds for plasma and CSF p-tau217 were set at 2 SD from the mean of CU PREVENT-AD Aβ-PET negative individuals (plasma p-tau217: 3.98 pg/mL; CSF p-tau217: 400.19 pg/mL).^17^ **eTables 8-13** further show the sensitivity, specificity, negative predictive value and positive predictive value of all other possible thresholds.

### Statistical analyses

Demographic and clinical variables by biomarker and by AT classification were compared using Kruskal-Wallis tests followed by Dunn’s post-hoc test for continuous variables and Fisher exact tests for categorical variables. Cox proportional hazard models were used to assess the risk of CI progression in by A/T groups and linear mixed models to assess longitudinal cognitive changes across the biomarker groups. We then tested the cognitive performance over time across the different biomarker groups using linear mixed-effects models and sex and education as potential confounders. Spearman’s rank test was used to evaluate the association between the biomarkers. Logistic regressions were used to test the performance of plasma, CSF and PET biomarkers as continuous variables in discriminating CI participants. Receiver operating characteristic (ROC) analyses and the resulting areas under the curve (AUCs) were also computed to assess the biomarker accuracy in distinguishing between CU and CI participants, alone or when combined. Model fits were compared using Vuong test. Two-sided p values < 0.05 were deemed significant. The analyses were performed using R programming language.

## Results

### Participants

A total of 62 participants (27%) developed CI during the study follow-up (31% in the subsample with all biomarkers). The mean age of the full sample at baseline was 63 years, 72 % were female and 39 % were *APOE* ε4 carriers (**Table 1**). The full sample was comparable to the 93 participants of interest for which we have all biomarkers. The breakdown of the different A/T biomarker demographic and clinical profiles in the full and subsamples is presented in **eTables 1-7**. The mean cognitive follow-up after plasma classification was 5.65 years (SD = 1.45, range 1.01 – 10.47), 5.57 years (SD = 1.48, range 1.00 – 10.00) after CSF classification, and 4.18 years (SD = 1.49, range 1.01 – 6.07) after PET classification. Restricting these range for the 93 participants gave similar follow-up length (plasma 5.85 years, SD 1.26, range 2.10 – 10.47; CSF 5.69 years, SD 1.18, range 2.08 – 9.74, PET 4.25 years, SD 1.47, range 1.11 – 6.07). The full cognitive follow-up length, which for most participants included evaluations prior to the biomarker assessment, was 7.63 years (SD = 1.94, range: 1.00 – 10.47) for the plasma subsample, 7.59 years (SD =1.89, range: 1.89-10.47) for the CSF subsample, 8.11 (SD = 1.76, range: 1 – 10.49) for the PET subsample and 8.04 years (SD 1.71, range 2.99 – 10.47) for the 93 participants with all biomarkers.

### A/T classification across biomarkers

Using Aβ_42/40_ and p-tau217 plasma biomarkers to classify participants, 9% (8/93) were classified as A+T+, 35% (33/93) as A+T-, 3% (3/93) as A-T+ and 53% (49/93) as A-T-. Using Aβ_42/40_ and p-tau217 CSF biomarkers, 15% (14/93) were classified as A+T+, 3% (3/93) as A+T-, 2% (2/93) as A-T+ and 80% (74/93) as A-T-. Finally, using PET biomarkers, 3% (3/93) were classified as A+T+, 30% (28/93) as A+T-, 1% (1/93) as A-T+ and 66% (61/93) as A-T-. These numbers were not statistically different to the proportion found in the full sample (Fisher’s exact p > 0.05) and it is also similar to what has been found in other PET cohorts. ^4,5^

### Rate of progression from CU to CI across A/T groups when defined using fluid versus neuroimaging biomarkers

Eighty-eight percent (7/8) of the A+T+_plasma_ group developed CI compared with 36% (12/33) in the A+T-_plasma_ group, 100% (3/3) in the A-T+ _plasma_ group and 14% (7/49) in the A-T-_plasma_ group (**Figure 2A-C**). The proportion of CU developing CI was higher in the A+T+ group when compared with A-T-_plasma_ and A+T-_plasma_ groups (p < 0.001, p = 0.01). When the groups were classified using CSF, 86% (12/14) of the A+T+ _CSF_, 33% (1/3) of the A+T-_CSF_ group, 50% (1/2) of the A-T+ and 20% (15/74) of the A-T-_CSF_ group developed CI (**Figure 2B**). An increased CU to CI progression rate was found in the A+T+ _CSF_ group when compared A-T-_CSF_, but no differences were found when compared to A+T-_CSF_ group (Fisher’s exact p < 0.001 and p = 0.12 respectively). In the PET groups, 100% (3/3) of A+T+ _PET_ biomarker group, 46% (13/28) of the A+T-_PET_ group, 100% (1/1) of the A-T+ and 20% (12/61) of the A-T-_PET_ group progressed to MCI (**Figure 2C**). The A+T+ _PET_ and A+T-_PET_ groups were associated with increased progression to CI when compared with A-T- _PET_ (Fisher’s exact p = 0.01, p = 0.01 respectively). Results were replicated when taking advantage of all available data, with all A+T+ groups showing a higher percentage of progression when compared to their respective A-T-groups (the % of CU who progressed to CI was 76 with plasma, 72 with CSF and 100% with PET in the A+T+ compared to 18-21% in the A-T-groups, **eFigure 2** in Supplement).

**Figure 2.**
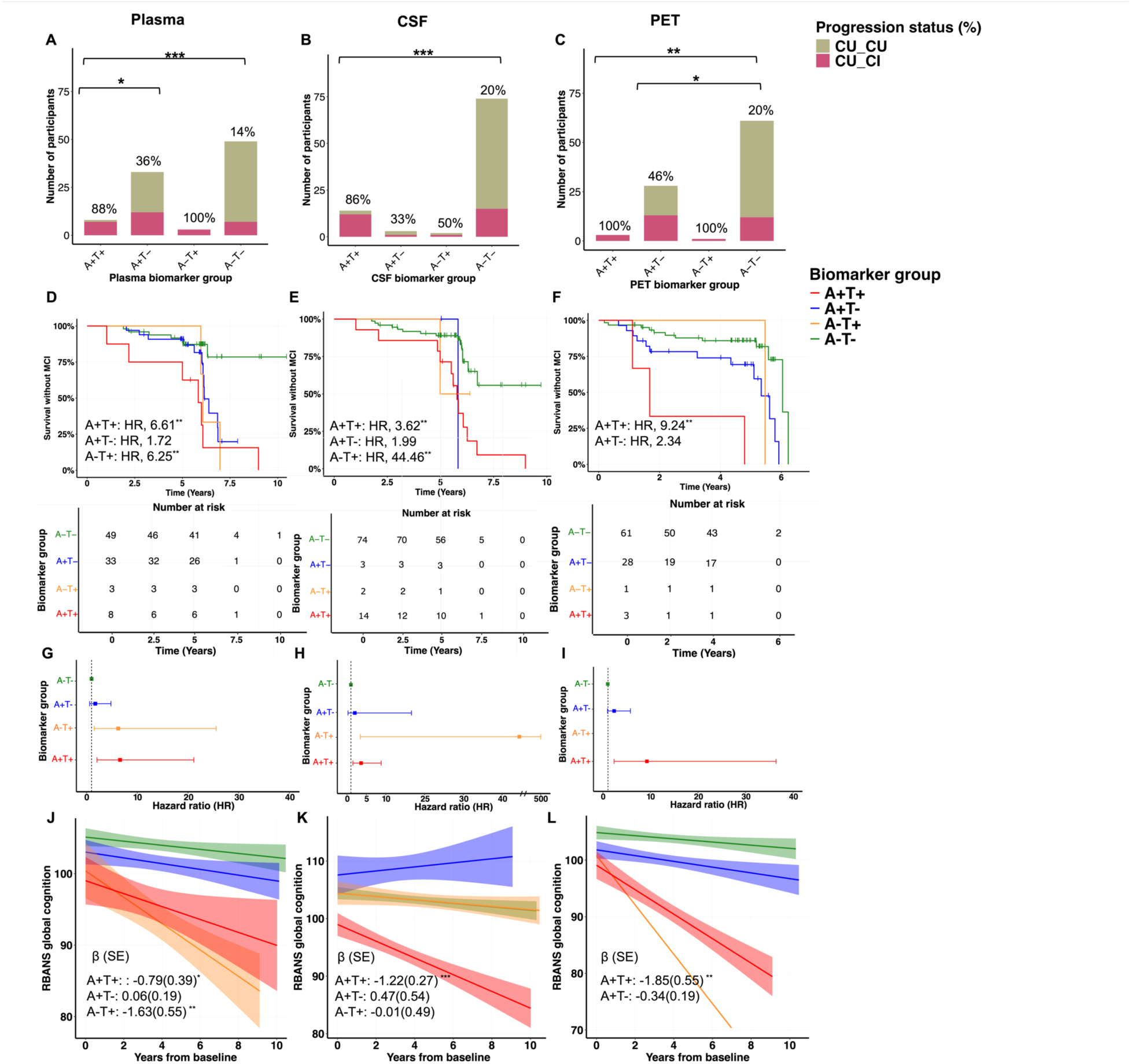
Clinical Progression to Cognitive Impairment (CI) across plasma, CSF and PET AT biomarker groups. **A-C)** Bar graphs represent the proportion of participants who developed CI across A) plasma Aβ_42/40_ and p-tau217; B) CSF Aβ_42/40_ and p-tau217; and C) PET biomarker profiles measured with ^18^F-NAV4694 and ^18^F-Flortaucipir. **D-F)**. Survival curves reflecting the progression to CI across D) plasma, E) CSF and F) PET biomarker groups. The vertical ticks on the curves refer to the censored participants, i.e., the loss of follow-up of the individuals. (G-I) Forest plots showing HR and 95% confidence intervals from the survival analyses. (J-L) Linear mixed effects models show the total cognitive score of RBANS over time across J) plasma, K) CSF and L) PET biomarker profiles. The linear mixed effects models analyses included annual cognitive data before and following plasma, CSF, and PET measures. Models included age at baseline, sex, and years of education as covariates. *Notes*: CU_CU = cognitively unimpaired older adults at the time of the biomarker measurement and remained cognitively unimpaired during follow-up; CU_CI = cognitively unimpaired older adults at the time of the biomarker measurement, who progressed to cognitive impairment during follow-up. The A-T- group was used as reference. The A-T+ in the PET biomarker group is displayed for visualization purposes but was not included in the statistical analyses. HR = hazard ratios; * *P* < 0.05*, ** P* < 0.01*, *** P* < 0.001.

One hundred twenty-eight of these 155 PET participants were included in a previously published study ^4^ that showed that 55% of the participants classified as A+T+_PET_, 9% of A+T-_PET_ and 9% of A-T-_PET_ developed CI when followed for a mean of 3.16 years after the A/T classification. All (100%) A+T+, 42% of A+T-_PET_ and 19% of the A-T-_PET_ groups have now developed CI after an additional 2.4 year of cognitive follow-up (**eFigure 3** in Supplement).

**Figure 3.**
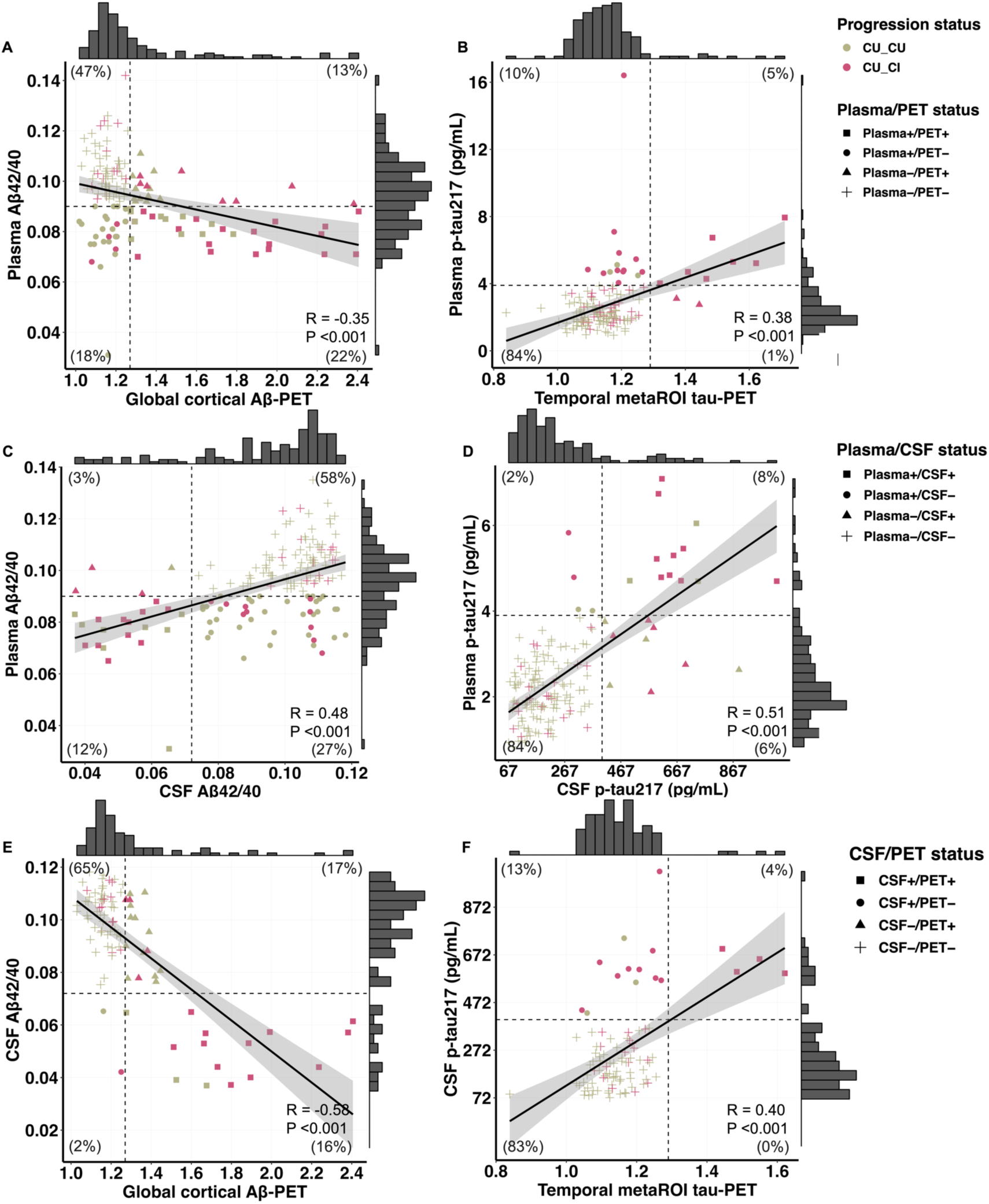
Scatterplots reflecting concordance status between plasma, CSF and PET Aβ and tau biomarkers. Correlation plots of A) plasma Aβ_42/40_ vs Aβ-PET biomarkers; B). plasma p-tau217 vs metaROI tau-PET SUVR values; C) plasma Aβ_42/40_ vs CSF Aβ_42/40_ biomarkers; D) plasma p-tau217 vs CSF p-tau217 biomarkers; E) CSF Aβ_42/40_ vs Aβ-PET biomarkers; F) CSF p-tau217 vs tau-PET biomarkers. Colors indicate the cognitive status of participants and symbols indicate the status of participants as being Aβ/tau positive or negative, vertical and horizontal dashed lines correspond to Plasma, PET and CSF biomarker cutoff values, respectively. *Notes*: we used 0.09 for plasma Aβ_42/40_ ; 0.072 for CSF Aβ_42/40_ ;1.27 SUVR for Aβ-PET; 3.98 for plasma p-tau217; 400.19 for CSF p-tau217; and 1.29 for tau-PET positivity. The total number of participants was n= 143 for individuals with both plasma and PET measurements, n = 158 for participants with both plasma and CSF measurements and n = 93 for participants with both CSF and PET.

Cox proportional hazard models showed a higher risk of progression from CU to CI among the A+T+_plasma_ and A-T+ _plasma_ (hazard ratios (HR) = 6.61, p = 0.001, 95%CI = 2.06 - 21.17; HR = 6.25; p = 0.01, 95%CI = 1.53-25.54; model concordance value (model fit) = 0.69; SE = 0.07; **Figure 2D & G**) when compared to A-T-_plasma_ (reference) group. In the CSF sample, we found an increased in the risk among A+T+_CSF_ and A-T+_CSF_ (HR = 3.62, p = 0.005, 95%CI = 1.49 – 8.81; HR = 44.46, p = 0.004, 95%CI = 3.44 – 573.91; model concordance value = 0.71; SE = 0.07; **Figure 2E & H)** compared to A-T-_CSF_ group. Finally, A+T+_PET_ participants exhibited a higher risk of CI progression compared to A-T-_PET_ (HR = 9.24, p = 0.001, 95% CI = 2.34 – 36.43; model concordance value = 0.70; SE = 0.06; **Figure 2F & I**). The A-T+ group was not included in the analyses given that only one participant was classified as A-T+ _PET_, this participant nevertheless developed CI during the study follow-up. All results were replicated in the larger sub-cohorts and giving the increased number of participants, the A+T-_PET_ now showed an increased risk of progression when compared to A-T-_PET_ group (HR = 2.75, p = 0.002, 95% CI = 1.43 – 5.27).

### Cognitive trajectories

We also investigated the longitudinal cognitive performance of participants within the AT biomarker groups while taking advantage of all cognitive time points, including the ones before the biomarker classifications were available. The A+T+_plasma_ and A-T+_plasma_ groups demonstrated a steeper cognitive decline compared to A-T-_plasma_ (reference) group (β = -0.79, p = 0.04, SE = 0.39, %95CI = -1.56 – -0.02; β = -1.63, p = 0.004, SE =0.55, 95% CI = -2.72 – -0.54; R^2^ = 0.08; **Figure 2J)** while no differences was observed between the A-T-_plasma_ and A+T-_plasma_ group (β = 0.06, p = 0.74, SE = 0.19, %95CI = -0.32 – 0.46). The A+T+_CSF_ group showed faster decline over time compared to A-T-_CSF_ (β = -1.22, p < 0.001, SE = 0.27, 95%CI = -1.75 – -0.70; R^2^ = 0.13, **Figure 2K**), but no difference was found between the reference group and A+T-_CSF_ nor the A-T+_CSF_ groups (β = 0.47, p = 0.38, SE = 0.54, 95% CI = -0.59 – 1.54; β = -0.01, p = 0.97, SE = 0.49, 95%CI = -1.00 – 0.97). The A+T+_PET_ group demonstrated a steeper cognitive decline compared to A-T-_PET_ (β = -1.85, p = 0.001, SE = 0.55, 95%CI = -2.94 – -0.76; **Figure 2L**). The A+T^_^_PET_ group demonstrated no differences when compared to A-T-_PET_ group (β = -0.34, p = 0.09, SE = 0.19, 95%CI = -0.73 – 0.06). Identical results were found in the full sample (see Supplement for more details).

### Concordance between different biomarkers modalities

We found weak correlation between plasma Aβ_42/40_ and Aβ -PET (R = -0.35, p < 0.001, **Figure 3A**), and between plasma p-tau217 and tau-PET (R = 0.38, p < 0.001, **Figure 3B**), but moderate correlations between plasma and CSF Aβ_42/40_ (R = 0.48, p < 0.001, **Figure 3C**) and plasma and CSF p-tau217 (R = 0.51, p < 0.001, **Figure 3D**). Similarly, CSF Aβ_42/40_ and CSF p-tau217 were moderately correlated with both Aβ and tau-PET (R = -0.58; R = 0.40; p < 0.001 respectively, **Figure 3E&F**). See **eFigure 4** supplementary results for the concordance when stratified by +/- status and for the percentage of participants who progressed to CI by biomarkers status.

**Figure 4.**
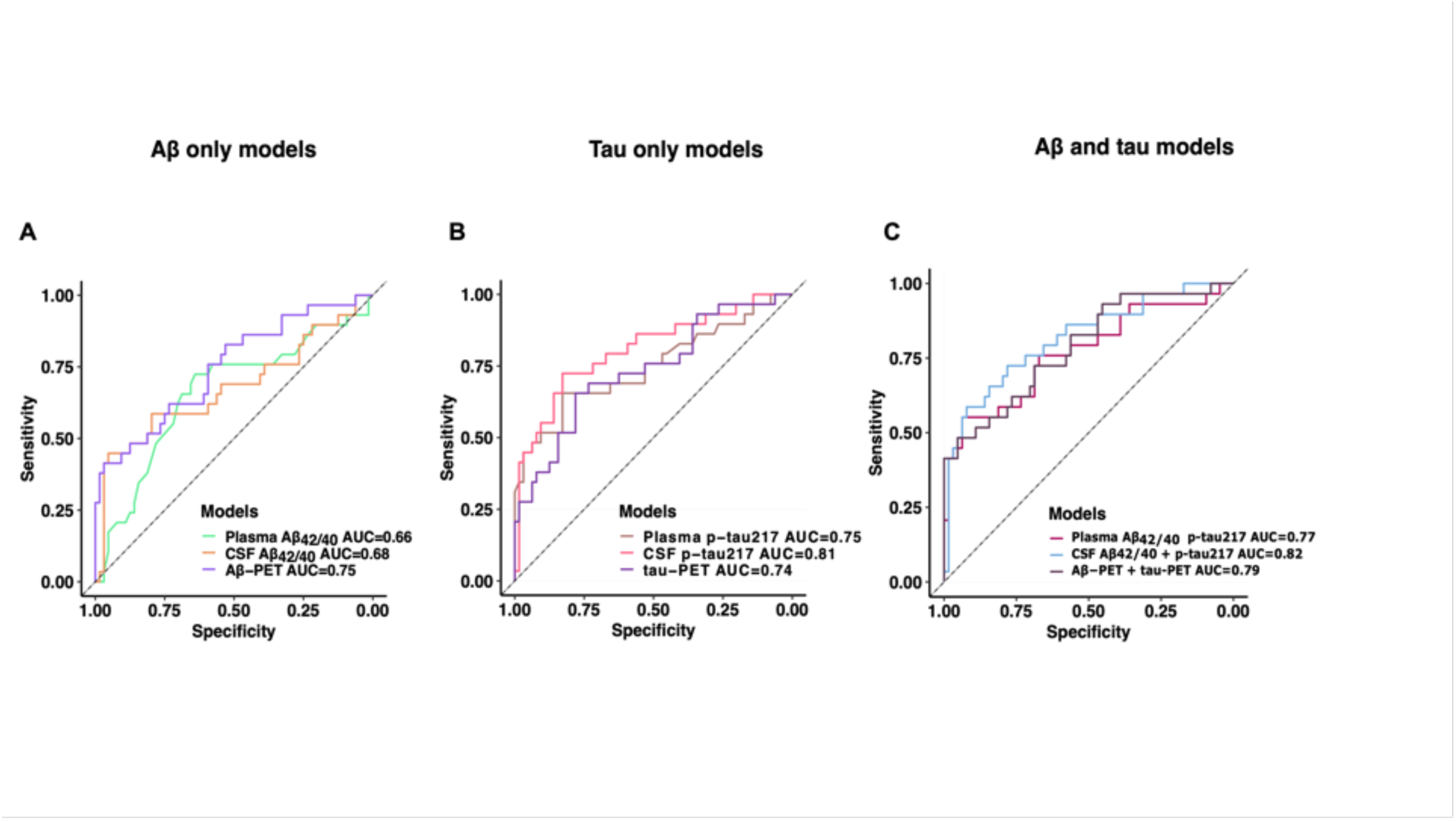
Discriminative accuracy of Plasma, CSF and PET biomarkers for identifying individuals who will develop cognitive impairment later. Receiver Operating Characteristic (ROC) curves and corresponding areas under the curve (AUC) showing the discriminative ability of A) individual plasma, CSF, and PET Aβ models; B) individual plasma, CSF, and PET tau models and C) the combination of plasma Aβ_42/40_,CSF Aβ_42/40_ and Aβ-PET biomarkers with plasma p-tau217, CSF p-tau217 and tau-PET in distinguishing between individuals who remained cognitively normal versus those who develop cognitive impairment.

### Direct comparisons between fluid and imaging biomarkers

All ROC curve predictive models were similar when compared using Delong test, with AUC ranging from .66 to .75 for Aβ, from .74 - .75 for tau and between .77 to .82 for their combinations (**Figure 4)**. Comparing the performance of Aβ models, Aβ-PET showed the best model fit when compared to plasma and CSF Aβ_42/40_ models (p < 0.05). Tau models and the combination of tau + Aβ models were comparable using Vuong test.

Finally, the biomarker cutoffs used in **Figure 1** gave good to excellent specificity (97% for plasma, 95% for CSF and 100% for PET), but low sensitivity (34%, 45% and 17% respectively) at identifying CU who will develop CI. Except for CSF, Aβ biomarkers cutoffs had low specificity (66% for plasma, 94% for CSF and 77% for PET), and low sensitivity (65%, 45% and 52% respectively).

## Discussion

Is still a matter of debate if CU individuals with Aβ and tau are condemned to progress to clinical AD. We found that all individuals with abnormal tau PET values, and almost all individuals with plasma p-tau217 values (between 75-90% depending on the sample) developed CI during a 10-year follow-up. When individuals were classified based on Aβ and tau biomarkers, higher clinical progression rate was found for all A+T+ groups compared to their respective A+T- group, with HRs up to 6.61, 3.62 and 9.24 for plasma, CSF, and PET respectively. Plasma biomarkers had the main advantages over PET of identifying about five time more T+ individuals. While these results suggest that p-tau217 can be used as a stand-alone test to identify CU who will develop CI, plasma biomarkers are however known to be more prone to measurement errors, matrix effects, and batches to batches variability, ^18,19^ limitations that will need to be considered before implementing plasma p-tau217 into memory clinics. Given the possible distress cause by the disclosure of such results, we also strongly advocate for restricting the use of plasma p-tau217 to specialized clinics with available counseling until a drug is approved for preclinical AD.

PET biomarkers are expensive and not widely available. Robust and accurate blood-based markers for AD are needed for clinical evaluation, trial recruitment and to identify individuals who could benefit from disease modifying therapies.^20–23^ Plasma p-tau217 has been found to differentiate CU from individuals with clinical AD, ^24^ it can detect AD pathology in individuals with mild CI, ^25^ and it correlates with Aβ PET, tau PET and cognitive decline in CU participants. ^26–30^ Our prospective study shows that plasma p-tau217 can also be used to predict the development of CI in CU individuals’ years before cognitive onset.

Aβ did not improve the predictive value of the tau biomarkers, which can mainly be explained by the fact that almost all T+ individuals were also A+. As for individuals abnormal Aβ but normal tau biomarkers, their risk of progression to CI was increased when classified with PET in comparison to the A-T-_PET_ group. Furthermore, 128 participants from this study were included in a previous publication in which only 9% of the A+T- participant had developed CI at the time,^4^ from these A+T- individuals 42% have now developed CU, suggesting that A+T- were just further away on the AD spectrum.

The main limitation of this study, which is also its main strength, is that unimpaired vs impaired cognitive classification was done blind to *APOE* status, MRI and CSF and PET biomarkers results. Not all CI individuals are therefore on the path towards AD, which explains why some A-T- individuals developed CI. ^31^ One other limitation is the low sample size in the A+T+ groups, which was nevertheless similar to what was found in previous PET studies. ^4,5^ The low racial and ethnical diversity of our population should also be noted as the findings may not be applicable to populations with greater ethnic diversity. The collection of plasma and CSF markers also often preceded PET measurements by a year or two, which prevent us from drawing strong conclusions about which biomarker predict a more imminent risk of progression. Finally, one contentious point while dichotomizing biomarkers is that various cutoff strategies might produce different results. This dichotomization is however needed to guide treatment decision.

In this longitudinal multimodal biomarker study spanning more than 10 years, 27% developed CI based on a multidisciplinary classification consensus meeting blind to biomarkers and *APOE* genotype information. While not interchangeable, fluid and PET biomarkers of AD pathology are both extremely valuable at identifying individuals who will develop CI with almost all individuals with p-tau217 abnormal values developing CI within a 10-year follow-up.

## Acknowledgements

**Funding/Support** The authors acknowledge all the PREVENT-AD participants and their families as well as all the PREVENT-AD team members for their time and dedication. The authors would like to acknowledge Alfonso Fajardo, Ting Qiu, Mohammadali Javanray, Jonathan Gallego Rudolf, Bery Mohammediyan, Jordana Remz for providing advice on the study analyses and **J**ennifer Tremblay-Mercier, Louise Hudon, Christine Dery, Elisabeth Sylvian, Gabriel Jean, Nolan-Patrick Conningham for their contribution to the data collection. Lobna Almasalmeh and the Neurokemi lab at Gothenburg university for their assistance in plasma and CSF sample processing. The project was funded by the Canadian Institute of Health Research (CIHR) (#438655) and Brain Canada grants. Dr. Zetterberg is a Wallenberg Scholar and a Distinguished Professor at the Swedish Research Council supported by grants from the Swedish Research Council (#2023-00356; #2022- 01018 and #2019-02397), the European Union’s Horizon Europe research and innovation programme under grant agreement No 101053962, Swedish State Support for Clinical Research (#ALFGBG-71320), the Alzheimer Drug Discovery Foundation (ADDF), USA (#201809- 2016862), the AD Strategic Fund and the Alzheimer’s Association (#ADSF-21-831376-C, #ADSF-21-831381-C, #ADSF-21-831377-C, and #ADSF-24-1284328-C), the Bluefield Project, Cure Alzheimer’s Fund, the Olav Thon Foundation, the Erling-Persson Family Foundation, Familjen Rönströms Stiftelse, Stiftelsen för Gamla Tjänarinnor, Hjärnfonden, Sweden (#FO2022- 0270), the European Union’s Horizon 2020 research and innovation programme under the Marie Skłodowska-Curie grant agreement No 860197 (MIRIADE), the European Union Joint Programme – Neurodegenerative Disease Research (JPND2021-00694), the National Institute for Health and Care Research University College London Hospitals Biomedical Research Centre, and the UK Dementia Research Institute at UCL (UKDRI-1003).

Frédéric St-Onge was funded by a scholarship from the Fonds de Recherche du Quebec – Santé (FRQS). Dr. Soucy is funded by the CIHR, Brain Canada, and Biogen Canada. Dr. Poirier is funded by CIHR, the J.L. Levesque Foundation, FRQS, and Natural Sciences and Engineering Research Council of Canada (NSERC) grants. MS receives funding from the Knut and Alice Wallenberg Foundation (Wallenberg Centre for Molecular and Translational Medicine; KAW2014.0363 and KAW 2023.0371), the Swedish Research Council (2017-02869, 2021-02678, 2021-06545 and 2023-06188), the European Union’s Horizon Europe research and innovation program under grant agreement no 101132933 (AD-RIDDLE) and 101112145 (PROMINENT), the National Institute of Health (R01 AG081394-01), the Swedish state under the agreement between the Swedish government and the County Councils, the ALF-agreement (ALFGBG- 813971 and ALFGBG-965326), the Swedish Brain Foundation (FO2021-0311), the Swedish Alzheimer Foundation (AF-994900), the Sahlgrenska Academy at the University of Gothenburg, the Västra Götaland Region R&D (VGFOUREG-995510) and Innovation platforms, Sahlgrenska Science Park and the National Institute for Health and Care Research University College London Hospitals Biomedical Research Centre.

## Conflict of Interest Disclosures

HZ has served at scientific advisory boards and/or as a consultant for Abbvie, Acumen, Alector, Alzinova, ALZPath, Amylyx, Annexon, Apellis, Artery Therapeutics, AZTherapies, Cognito Therapeutics, CogRx, Denali, Eisai, Merry Life, Nervgen, Novo Nordisk, Optoceutics, Passage Bio, Pinteon Therapeutics, Prothena, Red Abbey Labs, reMYND, Roche, Samumed, Siemens Healthineers, Triplet Therapeutics, and Wave, has given lectures in symposia sponsored by Alzecure, Biogen, Cellectricon, Fujirebio, Lilly, Novo Nordisk, and Roche, and is a co-founder of Brain Biomarker Solutions in Gothenburg AB (BBS), which is a part of the GU Ventures Incubator Program (outside submitted work).MS has served on advisory boards for Roche, Novo Nordisk and Servier, received speaker honoraria from Bioarctic, Eisai, Genentech, Novo Nordisk and Roche and receives research support (to the institution) from Alzpath, Bioarctic, Novo Nordisk and Roche (outside scope of submitted work). He is a co-founder of Centile Bioscience Ltd. No other disclosures were reported.

## Data availability

Data used in the preparation of this manuscript were obtained from the Pre-symptomatic Evaluation of Experimental or Novel Treatments for Alzheimer’s Disease (PREVENT-AD). Some of the data are publicly available (https://openpreventad.loris.ca and https://registeredpreventad.loris.ca), and the remaining data can be shared upon approval by the scientific committee at the Centre for Studies on Prevention of Alzheimer’s Disease (StoP-AD) at the Douglas Mental Health University Institute. A complete listing of PREVENT-AD investigators can be found at https://preventad.loris.ca/acknowledgements/acknowledgements.php?date=2023-03-23.

## Code availability

The code used for the statistical analyses is available from the first author upon request. AThe analyses and figures were built using R programming language (v.4.2.2) and R studio “Elsbeth Geranium” Release (7d165dcfc1b6d300eb247738db2c7076234f6ef0, 2022-12-03) for macOS (Packages: survival v3.4-0; survminer v0.4.9; lme4 v1.1-31; lmerTest v3.1-3; ggplot v2.3.5.0; tidyverse v1.3.2; dplyr v1.1.2; stats v4.2.1; sjPlot v2.8.12; zoo v1.8-12; tibble v3.2.1; rstatix v0.7.2; pROC v1.18.4).

## Contributions

Yara Yakoub, Nicholas J. Ashton, Michael Schöll, Pedro Rosa-Neto, Judes Poirier, John C. S. Breitner, Henrik Zetterberg, Kaj Blennow, and Sylvia Villeneuve contributed to the study concept and design. Yara Yakoub, Nicholas J. Ashton, Thomas K. Karikari, Christine Dery, Frédéric St-Onge, Maiya Geddes, Simon Ducharme, Maxime Montembeault, and Jean-Paul Soucy contributed to data acquisition and analysis. Yara Yakoub and Sylvia Villeneuve drafted the manuscript and figures.

## Notes

### Author Declarations

Written informed consent was obtained from all participants and all research procedures were approved by the Institutional Review Board at McGill University and complied with the ethical principles of the Declaration of Helsinki. A detailed description of the PREVENT-AD cohort is available (https://pubmed.ncbi.nlm.nih.gov/34192666/)

## References

1. Jack CR, Jr., Holtzman DM. Biomarker modeling of Alzheimer’s disease. Neuron. Dec 18 2013;80(6):1347–58. doi:10.1016/j.neuron.2013.12.003

2. Association As. NIA-AA Revised Clinical Criteria for Alzheimer’s Disease. Alzheimer Dis Assoc Disord. 2023;

3. Jack CR, Bennett DA, Blennow K, et al. NIA-AA Research Framework: Toward a biological definition of Alzheimer’s disease. Alzheimer’s & Dementia: The Journal of the Alzheimer’s Association. 2018;14(4):535–562. doi:10.1016/j.jalz.2018.02.018

4. Strikwerda-Brown C, Hobbs DA, Gonneaud J, et al. Association of Elevated Amyloid and Tau Positron Emission Tomography Signal With Near-Term Development of Alzheimer Disease Symptoms in Older Adults Without Cognitive Impairment. JAMA Neurology. 2022;doi:10.1001/jamaneurol.2022.2379

5. Ossenkoppele R, Pichet Binette A, Groot C, et al. Amyloid and tau PET-positive cognitively unimpaired individuals are at high risk for future cognitive decline. Nature Medicine. 2022;28(11):2381–2387. doi:10.1038/s41591-022-02049-x

6. Villemagne VL, Lopresti BJ, Doré V, et al. What Is T+? A Gordian Knot of Tracers, Thresholds, and Topographies. Journal of Nuclear Medicine. 2021;62(5):614–619. doi:10.2967/jnumed.120.245423

7. Barthélemy NR, Li Y, Joseph-Mathurin N, et al. A soluble phosphorylated tau signature links tau, amyloid and the evolution of stages of dominantly inherited Alzheimer’s disease. Nat Med. Mar 2020;26(3):398–407. doi:10.1038/s41591-020-0781-z

8. Montoliu-Gaya L, Benedet AL, Tissot C, et al. Mass spectrometric simultaneous quantification of tau species in plasma shows differential associations with amyloid and tau pathologies. Nat Aging. Jun 2023;3(6):661–669. doi:10.1038/s43587-023-00405-1

9. Verberk IMW, Slot RE, Verfaillie SCJ, et al. Plasma Amyloid as Prescreener for the Earliest Alzheimer Pathological Changes. Ann Neurol. Nov 2018;84(5):648–658. doi:10.1002/ana.25334

10. Sperling RA, Sperling R, Donohue MC, et al. Trial of Solanezumab in Preclinical Alzheimer’s Disease. The New England journal of medicine. 2023;389(12):1096–1107. doi:10.1056/NEJMoa2305032

11. van Dyck CH, Swanson CJ, Aisen P, et al. Lecanemab in Early Alzheimer’s Disease. The New England journal of medicine. 2023;388(1):9–21. doi:10.1056/NEJMoa2212948

12. Tremblay-Mercier J, Madjar C, Das S, et al. Open science datasets from PREVENT-AD, a longitudinal cohort of pre-symptomatic Alzheimer’s disease. NeuroImage Clinical. 2021;31:102733. doi:10.1016/j.nicl.2021.102733

13. Meyer P-F, Ashton NJ, Karikari TK, et al. Plasma p-tau231, p-tau181, PET Biomarkers, and Cognitive Change in Older Adults. Annals of neurology. 2022;91(4):548–560. doi:10.1002/ana.26308

14. Gobom J, Parnetti L, Rosa-Neto P, et al. Validation of the LUMIPULSE automated immunoassay for the measurement of core AD biomarkers in cerebrospinal fluid. Clin Chem Lab Med. Jan 27 2022;60(2):207–219. doi:10.1515/cclm-2021-0651

15. Gonzalez-Ortiz F, Ferreira PCL, González-Escalante A, et al. A novel ultrasensitive assay for plasma p-tau217: Performance in individuals with subjective cognitive decline and early Alzheimer’s disease. Alzheimers Dement. Feb 2024;20(2):1239–1249. doi:10.1002/alz.13525

16. Jack CR, Jr., Wiste HJ, Weigand SD, et al. Defining imaging biomarker cut points for brain aging and Alzheimer’s disease. Alzheimers Dement. Mar 2017;13(3):205–216. doi:10.1016/j.jalz.2016.08.005

17. Therriault J, Servaes S, Tissot C, et al. Equivalence of plasma p-tau217 with cerebrospinal fluid in the diagnosis of Alzheimer’s disease. Alzheimer’s & Dementia. 2023/11/01 2023;19(11):4967–4977. doi: 10.1002/alz.13026

18. Mattsson-Carlgren N, Palmqvist S, Blennow K, Hansson O. Increasing the reproducibility of fluid biomarker studies in neurodegenerative studies. Nat Commun. Dec 7 2020;11(1):6252. doi:10.1038/s41467-020-19957-6

19. Hansson O, Mikulskis A, Fagan AM, et al. The impact of preanalytical variables on measuring cerebrospinal fluid biomarkers for Alzheimer’s disease diagnosis: A review. Alzheimer’s & Dementia. 2018/10/01/ 2018;14(10):1313–1333. doi: 10.1016/j.jalz.2018.05.008

20. Hansson O, Blennow K, Zetterberg H, Dage J. Blood biomarkers for Alzheimer’s disease in clinical practice and trials. Nat Aging. May 2023;3(5):506–519. doi:10.1038/s43587-023-00403-3

21. Hampel H, Hu Y, Cummings J, et al. Blood-based biomarkers for Alzheimer’s disease: Current state and future use in a transformed global healthcare landscape. Neuron. 2023;111(18):2781–2799. doi:10.1016/j.neuron.2023.05.017

22. Mattsson-Carlgren N, Collij LE, Stomrud E, et al. Plasma Biomarker Strategy for Selecting Patients With Alzheimer Disease for Antiamyloid Immunotherapies. JAMA Neurol. Jan 1 2024;81(1):69–78. doi:10.1001/jamaneurol.2023.4596

23. Mattsson-Carlgren N, Salvadó G, Ashton NJ, et al. Prediction of Longitudinal Cognitive Decline in Preclinical Alzheimer Disease Using Plasma Biomarkers. JAMA Neurology. 2023;doi:10.1001/jamaneurol.2022.5272

24. Palmqvist S, Tideman P, Cullen N, et al. Prediction of future Alzheimer’s disease dementia using plasma phospho-tau combined with other accessible measures. Nature Medicine. 2021/06/01 2021;27(6):1034–1042. doi:10.1038/s41591-021-01348-z

25. Mattsson-Carlgren N, Janelidze S, Palmqvist S, et al. Longitudinal plasma p-tau217 is increased in early stages of Alzheimer’s disease. Brain. 2021;143(11):3234–3241. doi:10.1093/BRAIN/AWAA286

26. Jonaitis EM, Janelidze S, Cody KA, et al. Plasma phosphorylated tau 217 in preclinical Alzheimer’s disease. Brain Communications. 2023;5(2):fcad057. doi:10.1093/braincomms/fcad057

27. Doré V, Doecke JD, Saad ZS, et al. Plasma p217+tau versus NAV4694 amyloid and MK6240 tau PET across the Alzheimer’s continuum. Alzheimers Dement (Amst). 2022;14(1):e12307. doi:10.1002/dad2.12307

28. Rissman RA, Langford O, Raman R, et al. Plasma Aβ42/Aβ40 and phospho-tau217 concentration ratios increase the accuracy of amyloid PET classification in preclinical Alzheimer’s disease. Alzheimer’s & Dementia. 2024;20(2):1214–1224. doi: 10.1002/alz.13542

29. Barthélemy NR, Salvadó G, Schindler SE, et al. Highly accurate blood test for Alzheimer’s disease is similar or superior to clinical cerebrospinal fluid tests. Nature Medicine. 2024/02/21 2024;doi:10.1038/s41591-024-02869-z

30. Janelidze S, Berron D, Smith R, et al. Associations of Plasma Phospho-Tau217 Levels With Tau Positron Emission Tomography in Early Alzheimer Disease. JAMA Neurology. 2021;78(2):149–156. doi:10.1001/jamaneurol.2020.4201

31. Gauthier S, Reisberg B, Zaudig M, et al. Mild cognitive impairment. Lancet. Apr 15 2006;367(9518):1262–70. doi:10.1016/s0140-6736(06)68542-5

