## Supplement for "Plasma p-tau217 predicts cognitive impairments up to ten years before onset in normal older adults"

**Total word count of the main text in Supplement**: 1615

**The supplementary includes** 10 references, 13 Table and 4 Figures

Mme. Yakoub 

Dr. Gonzalez-Ortiz

Dr. Ashton 

Mme. Christine Déry

Dr. Strikwerda-Brown 

Dr. St-Onge

Dr. Ourry

Dr. Schöll 

Dr. Geddes

Dr. Ducharme

Dr. Montembeault

Dr. Rosa-Neto

Dr. Soucy

Dr. Breitner 

Dr. Zetterberg 

Dr. Blennow 

Dr. Poirier 

Dr. Villeneuve

**Table of contents**

**eMethods**

**eResults**

**eReferences**

**eTable 1. Demographic Characteristics of participants across AT plasma subsample.**

**eTable 2. Demographic Characteristics of participants across AT CSF subsample.**

**eTable 3. Demographic Characteristics of participants across AT PET subsample.**

**eTable 4. Sample Demographics across plasma, CSF and PET full sample.**

**eTable 5. Demographic Characteristics of participants across AT plasma full sample.**

**eTable 6. Demographic Characteristics of participants across AT CSF full sample.**

**eTable 7. Demographic Characteristics of participants across AT PET full sample.**

**eTable 8. Diagnostic accuracy of plasma p-tau217 in predicting CI**

**eTable 9. Diagnostic accuracy of plasma Aβ_42/40_ in predicting CI**

**eTable 10. Diagnostic accuracy of CSF p-tau217 in predicting CI**

**eTable 11. Diagnostic accuracy of CSF Aβ_42/40_ in predicting CI**

**eTable 12. Diagnostic accuracy of metaROI tau-PET in predicting CI**

**eTable 13. Diagnostic accuracy of Aβ -PET in predicting CI**

**eFigure 1. Distribution of Plasma, CSF, PET, and cognitive measurements**

**eFigure 2. Clinical Progression to Cognitive Impairment (CI) across the full plasma, CSF and PET AT biomarker groups.**

**eFigure 3. Comparison of the proportion of cognitively impaired (CI) in Cognitively Unimpaired (CU) individuals on PET: replication of 2021 study and follow-up.**

**eFigure 4 Percentage of cognitively unimpaired vs impaired individuals between plasma, CSF and PETAβ and tau biomarkers.**

**eMethods**

### PREVENT-AD cohort

### The PREVENT-AD cohort is an ongoing longitudinal observational study of older adults with a family history of sporadic AD. PREVENT-AD participants were enrolled between 2011 and 2017, were 60+ years old, or 55-59 if within 15 years of their youngest-affected relative’s age of onset and CU at enrollment.^1^ Information on *APOE ε4* status, age, sex, years of education were collected at entry into the program. Normal cognition at enrollment was based on a brief cognitive screening at study entry using the Clinical Dementia Rating (CDR) and Montreal Cognitive Assessment (MoCA). In a few cases of ambiguous CDR (0.5) or MoCA (26), participants were evaluated by a certified neuropsychologist with an extensive neuropsychological battery assessment. Participants were then followed annually using the Repeatable Battery for the Assessment of Neuropsychological Status (RBANS) and other cognitive tasks. ^1^ Individuals at baseline who performed below the norms for the RBANS were also evaluated by a certified neuropsychologist to confirm normal cognition. Information on peripheral blood was available at baseline and at up to four annual follow-up visits (2011-2017). CSF collection via lumbar puncture, when available, followed a similar timeline. PET was introduced in 2017.

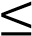

### Clinical classification

### All CI participants included in this study were classified as having memory problems with or without other cognitive deficit except for one participant in the plasma and PET sample who was classified as having isolated visuospatial impairments. When the CI classification was reviewed at more than one consensus meeting, the latest classification was used. Three individuals reverted to CU during the follow-ups; these individuals were considered as CU in all analyses.

**Plasma measures**

In the plasma sample, Aβ40 and Aβ42 concentrations were measured using ultrasensitive with immunoprecipitation coupled with mass spectrometry (IP-MS) technique using a KingFisher Flex Purification System (Thermo Fisher Scientific).^2^ Inter-assay coefficients of variation were <5%.^2^ Plasma p-tau217 concentrations were measured using an in-house Simoa platform developed at the Clinical Neurochemistry Laboratory, University of Gothenburg.^3^ Prior to analysis, the plasma samples were thawed, vortexed, and centrifuged (4000g for 10 min at room temperature). They were then examined using an HD-X analyzer, with identical reagent batches used throughout the study. To ensure quality control, three duplicate samples were included at the beginning and end of each run, resulting in an overall coefficient of variation of 7.9% for the biomarker measurement.

**Cerebrospinal Fluid (CSF)**

A total of 159 PREVENT-AD participants underwent a lumbar puncture (LP) procedure after fasting overnight. A Sprotte 24-gauge atraumatic needle was used. Approximately of 20–30 mL of samples were aliquoted (500 μL) into propylene cryotubes and stored at –80°C. ^4^ P-tau217 measured using an in-house Simoa platform developed at the Clinical Neurochemistry Laboratory, University of Gothenburg.^3^and CSF Aβ1-42, Aβ1-40 measured using Lumipulse G1200 fully automated immunoassay. ^5^

**PET preprocessing**

The PET scans were performed at the McConnell Brain Imaging Centre of the Montreal Neurological Institute, Canada. Aβ-PET images using ^18^F-NAV4694 as tracer were captured 40-70 minutes after injecting a targeted dose of 220 MBq (6 mCi). Tau-PET images using ^18^F-flortaucipir as tracer were obtained 80-100 minutes after injection with a targeted dose of 370 MBq (10 mCi). We acquired 4 frames of 5 minutes each. A transmission attenuation correction scan was also performed. The images were reconstructed using a three-dimensional (3D) ordinary Poisson ordered subset expectation maximum ([OP-OSEM], with 10 iterations and 16 subsets). Decay and motion corrections were applied to the images, and scatter correction was done using a 3D scatter estimation method.^6^ The MRI scan closest in time to the PET scan for each participant was selected for PET image preprocessing. The T1-weighted MRI images were preprocessed and divided into 34 bilateral regions of interest (ROI) based on the Desikan Killiany atlas using FreeSurfer v.5.3. ^7^ The PET images were realigned, temporally averaged, and co-registered to the closest T1-weighted image. Then, they were masked to remove CSF signal and smoothed using a 6-mm^3^ Gaussian kernel. Standardized Uptake Value Ratios (SUVRs) were calculated as the ratio of tracer uptake in the regions of interest compared to the uptake in the gray matter of the cerebellum for Aβ-PET scans or compared to the inferior cerebellar gray matter for tau-PET scans.^8,9^ An in-house pipeline (https://github.com/villeneuvelab/vlpp) was used for the preprocessing of all PET scans. The PET data underwent a new reconstruction when compared to the Strikwerda-Brown et al. study published in 2022.^10^ The PET sample overlaps with the former study, with addition to twenty-seven new participants, all of whom had scans taken between 2018 and 2021.

**eResults**

**Rate of progression from CU to CI across A/T groups when defined using fluid versus neuroimaging biomarkers.**

We replicated the same analyses using the full sample in each plasma, CSF, and PET. In the plasma sample, the results showed that, 76% (16/21) of the A+T+_plasma_ group developed CI compared to 23% (14/60) in the A+T- _plasma_ group, 71% (5/7) in the A-T+ _plasma_ group and 21% (25/127) in the A-T- _plasma_ group (e**Figure 2A-C**). Fisher’s exact test showed that the proportion of CU developing CI was higher in the A+T+ group when compared with A-T- _plasma_ and A+T- _plasma_ groups (Fisher’s exact p < 0.001, p = 0.01 respectively). The proportion of CI was also higher in the A+T- _plasma_ was also higher than A-T+ _plasma_ (Fisher’s exact p = 0.02).

When the groups were classified using CSF, 72% (13/18) of the A+T+ _CSF_, 20% (1/5) of the A+T- _CSF_ group, 33% (1/3) of the A-T+ and 18 % (24/133) of the A-T- _CSF_ group developed CI (e**Figure 2B**). An increased CU to CI progression rate was found in the A+T+ _CSF_ group when compared A-T- _CSF_ (Fisher’s exact p < 0.001), but no differences were found between the A+T- _CSF_ and A-T+_CSF_ groups when compared to A-T- _CSF_ reference group.

In the PET groups, 100% (8/8) of A+T+ _PET_ biomarker group, 44% (20/45) of the A+T- _PET_ group, 100% of the A-T+ (1/1) and 20% (20/101) of the A-T- _PET_ group developed CI (**eFigure 2C**). The A+T+ _PET_ group was associated with increased progression to CI when compared with A-T- _PET_ and A+T- _PET_ (Fisher’s exact p < 0.001, p = 0.005 respectively). We also found an increased risk of progression to CI in the A+T- _PET_ to A-T- _PET_ (Fisher’s exact p = 0.004)

Cox proportional hazard models showed a higher risk of progression from CU to CI among the A+T+_plasma_ and A-T+ _plasma_ (hazard ratios (HR) = 7.81, p < 0.001, 95%CI = 3.92 – 15.59; HR = 4.25; p = 0.004, 95%CI = 1.60 – 11.31; model concordance value = 0.72; SE **=** 0.042; e**Figure 2D**) when compared to A-T- _plasma_ (reference) group_._ There were no differences between A+T-_plasma_ and A-T-_plasma_ group (HR = 1.14, p = 0.69, 95%CI = 0.58 – 2.24). In the CSF sample, cox proportional hazard models showed an increased in the risk of CI progression among CU A+T+_CSF_ and A-T+_CSF_ (HR = 3.63, p < 0.001, 95%CI = 1.72 – 7.70; model concordance value = 0.68; SE **=** 0.06; e**Figure 2E)** compared to A-T-_CSF_ group. No differences were found in the A+T-_CSF_ nor A-T+_CSF_ when compared to A-T-_CSF_ reference group (HR = 1.50; *p* = 0.69, 95%CI = 0.19 – 11.69; HR = 3.82, p = 0.20, 95%CI = 0.50 – 29.24). Finally, A+T+_PET_ and A+T-_PET_ participants exhibited a higher risk of CI progression compared to A-T-_PET_ (HR = 9.30, p < 0.001*,* 95%CI = 3.67 – 23.55; HR = 2.75, p = 0.002, 95% CI = 1.43 – 5.27; model concordance value = 0.73; SE = 0.04; **eFigure 2F**). The A-T+ group was not included in the analyses given that only one participant was classified as A-T+_PET_, this participant nevertheless developed CI during the study follow-up.

We also investigated the longitudinal cognitive performance of participants within the AT biomarker groups using plasma, CSF and PET biomarkers while taking advantage of all cognitive time points, including the ones before the biomarkers’ classifications when available. The A+T+_plasma_ and A-T+_plasma_ groups demonstrated a steeper cognitive decline compared to A-T-_plasma_ (reference) group (β = -1.03, p < 0.001, SE =0.26, %95 CI = -1.53 – -0.52; β = -1.00, p = 0.02, SE = 0.42, 95% CI = -1.83 – -0.18; R^2^ = 0.12; **eFigure 2J)** while no differences was observed between the A-T-_plasma_ and A+T-_plasma_ group (β = 0.01, p = 0.96, SE =0.15, %95 CI = -0.29 – 0.31). In the CSF, the A+T+_CSF_ group showed a faster decline over time compared to A-T-_CSF_ (β = -1.24, p < 0.001, SE = 0.24, 95% CI = -1.72 – -0.77; R^2^ = 0.11; **eFigure 2K**), but no differences was found between the reference group and A+T-_CSF_ nor the A-T+_CSF_ groups (β = 0.13, p = 0.77, SE = 0.44, 95% CI = -0.74 – 1.00; β = 0.33, p = 0.48, SE = 0.47, 95%CI = -0.58 – 1.24). When the groups were classified based on PET, the A+T+_PET_ group demonstrated a steeper cognitive decline compared to A-T-_PET_ (β = -1.66, p < 0.001, SE = 0.38, 95%CI = -2.40 – -0.91; R^2^ = 0.11; **eFigure 2L**). The A+T^_^_PET_ group demonstrated no differences when compared to A-T_-PET_ group (β = - 0.27, p = 0.10, SE = 0.16, 95%CI = -0.59 – 0.05).

**Concordance between the different biomarkers.**

When these biomarkers were stratified by +/- status, Aβ plasma and PET biomarkers were concordant for 69% of the participants (22% Plasma+/PET+; 47% Plasma-/PET-) with a similar number of discrepancies between the Plasma-/PET+ (13%) and the Plasma+/PET- (18%). The concordance between plasma p-tau217 and tau PET was 89% (5% Plasma+/PET+; 84% Plasma-/PET-), with 1% Plasma-/PET+ and 10% Plasma+/PET-. The concordance between plasma and CSF Aβ_42/40_ was 70% (12% Plasma+/CSF+; 58% Plasma-/CSF-), with 3% Plasma-/CSF+ and 27% Plasma+/CSF-. The concordance between plasma and CSF p-tau217 was 92% (8% Plasma+/CSF+; 84% Plasma-/CSF-), with 6% CSF-/PET+ and 2% CSF+/PET-. The concordance between CSF and PET Aβ was 81% (16% Plasma+/CSF+; 65% Plasma-/CSF-), with 2% CSF-/PET+ and 17% CSF+/PET-. The concordance between CSF and PET tau was 87% (4% Plasma+/CSF+; 83% Plasma-/CSF-), with 0% CSF-/PET+ and 13% CSF+/PET-.

**eFigures & eTables**

**eFigure 1. Distribution of Plasma, CSF, PET, and cognitive measurements**

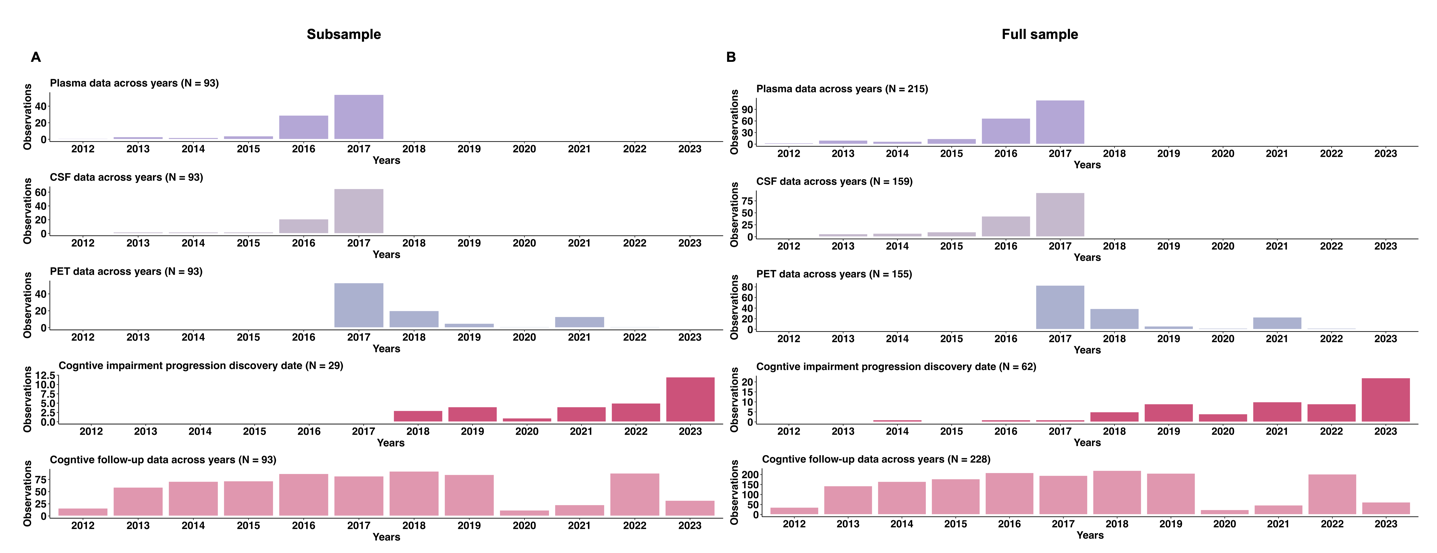

**eFigure 1.** Histograms illustrating the temporal distribution of assessments for plasma, CSF, and PET biomarkers, alongside the discovery dates of cognitive impairment progression, and longitudinal cognitive follow-up data across A). subsample and B). full sample.

**eFigure 2. Clinical Progression to Cognitive Impairment (CI) across the full plasma, CSF, and PET AT biomarker groups.**

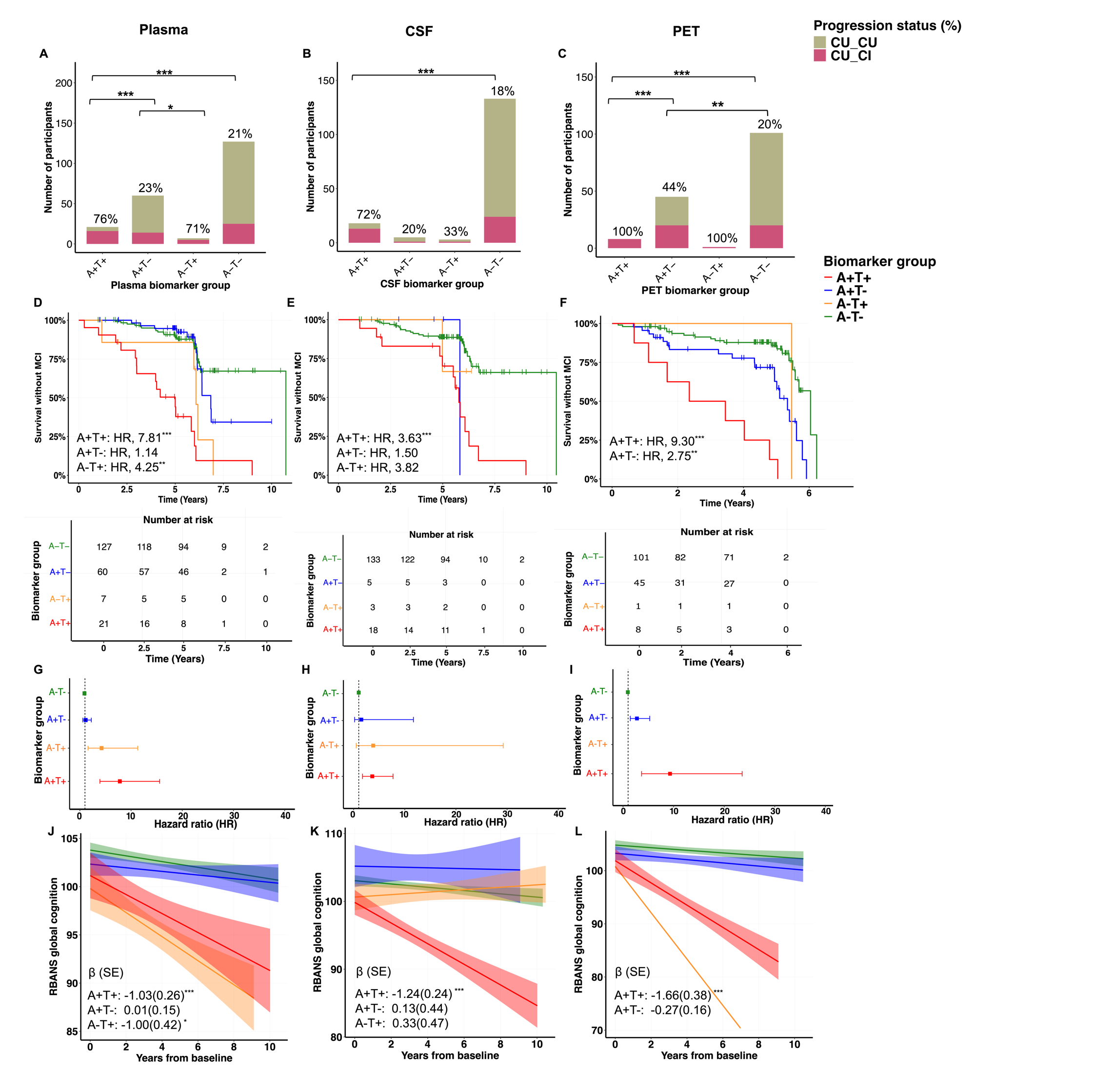

**eFigure 2. (A-C)** Bar graphs represent the proportion of participants who developed CI in the full sample across A) plasma Aβ_42/40_ and p-tau217; B) CSF Aβ_42/40_ and p-tau217; and C) PET biomarker profiles measured with ^18^F-NAV4694 and ^18^F-Flortaucipir. **C-E)**. Survival curves reflecting the progression to CI across C) plasma, D) CSF and E) PET biomarker groups. The vertical ticks on the curves refer to the censored participants, i.e., the loss of follow-up of the individuals. (G-I) Forest plots showing HR and 95% confidence intervals from the survival analyses. (J-L) Linear mixed effects models show the total cognitive score of RBANS over time across J) plasma, K) CSF and L) PET biomarker profiles. The linear mixed effects models analyses included annual cognitive data before and following plasma, CSF, and PET measures. Models included age at baseline, sex, and years of education as covariates. *Notes*: CU_CU = cognitively unimpaired older adults at the time of the biomarker measurement and remained cognitively unimpaired during follow-up; CU_CI = cognitively unimpaired older adults at the time of the biomarker measurement, who progressed to cognitive impairment during follow-up. The A-T- group was used as reference. The A-T+ in the PET biomarker group is displayed for visualization purposes but was not included in the statistical analyses*.* HR = hazard ratios*;* ^*^ *P* < 0.05*, ^**^ P* < 0.01*, ^***^ P* < 0.001*.*

**eFigure 3. Comparison of the proportion of cognitively impaired (CI) progressors in Cognitively Unimpaired (CU) individuals on PET: replication of 2021 study and follow-up.**

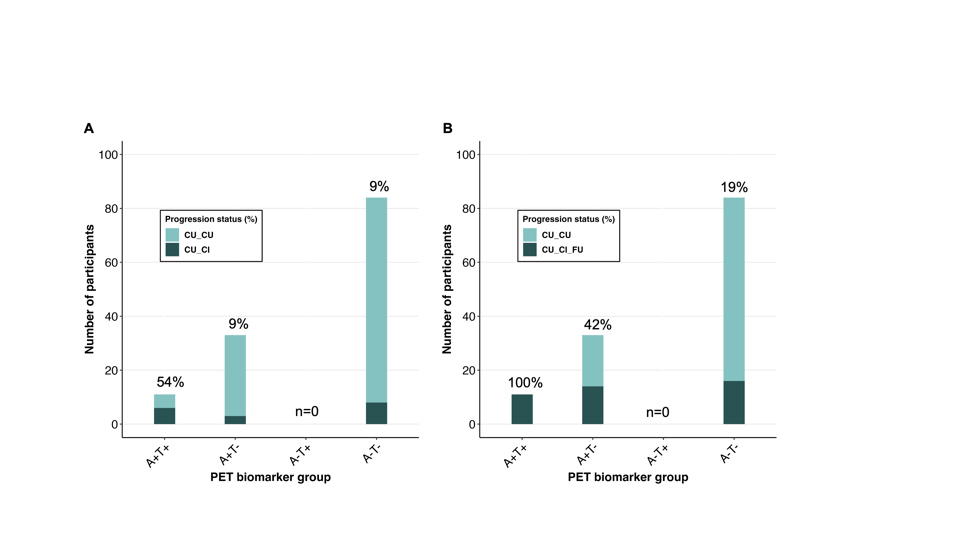

**eFigure 3.** Bar graphs comparing the proportion of cognitive impairment over a span of two years across 128 PET participants included in a previous publication. A). Distribution of cognitively impaired across AT PET biomarker profiles as reported in Strikwerda-Brown *JAMA Neurology* study in 2021; B) distribution of cognitively impaired across the same participants 2.4 years later. *Notes*: CU_CU: cognitively unimpaired older adults at the time of the biomarker measurement and remained cognitively unimpaired during follow-up; CU_CI: cognitively unimpaired older adults at the time of the biomarker measurement, who progressed to CI in 2021 study; CU_CI_FU: cognitively unimpaired older adults at the time of the biomarker measurement, who progressed to CI 2.4 years following the 2021 study.

**eFigure 4 Percentage of cognitively unimpaired vs impaired individuals between plasma, CSF and PETAβ and tau biomarkers.**

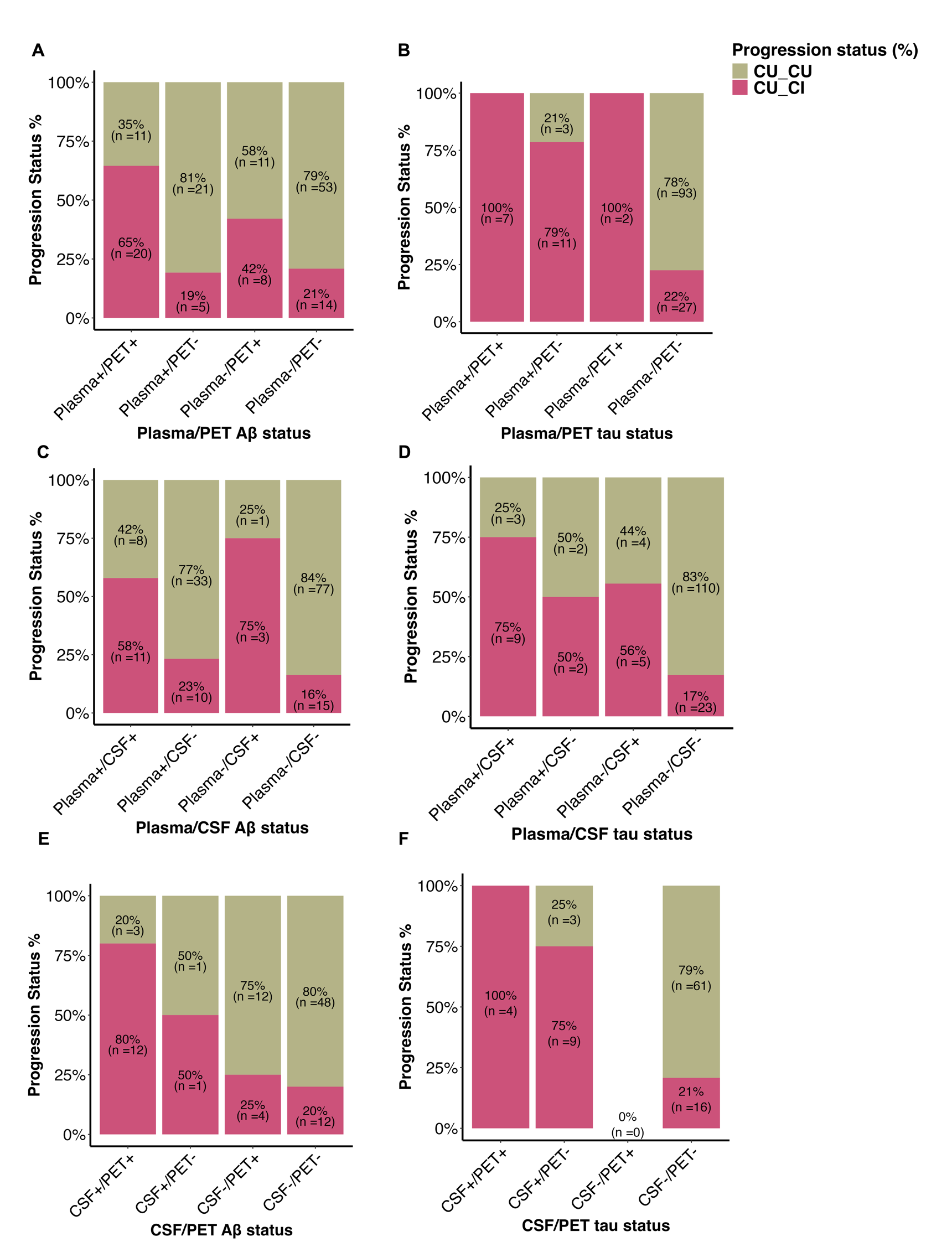

**eFigure 4.** Bar graphs showing the percentage of CU and CI across A) plasma Aβ_42/40_ vs Aβ-PET biomarkers; B). plasma p-tau217 vs metaROI tau-PET SUVR values; C) plasma Aβ_42/40_ vs CSF Aβ_42/40_ biomarkers; D) plasma p-tau217 vs p-tau217 biomarkers. Colors indicate the cognitive status of participants. *Notes*: we used 0.09 for plasma Aβ_42/40_ ; 0.072 for CSF Aβ_42/40_ ;1.27 SUVR for Aβ-PET; 3.98 for plasma p-tau217; 400.19 for CSF p-tau217; and 1.29 for tau-PET positivity. The total number of participants was n = 143 for individuals with both plasma and PET measurements, n = 158 for participants with both plasma and CSF measurements and n = 93 for participants with both CSF and PET.

**eTable 1. Demographic Characteristics of participants across AT plasma subsample.**

|  | **A+T+** | **A+T-** | **A-T+** | **A-T-** | **Group differences** |
| --- | --- | --- | --- | --- | --- |
|  | **(n = 8)** | **(n = 33)** | **(n = 3)** | **(n = 49)** |  |
| **Age at baseline, years** | 63.23 (4.76) | 63.82 (5.11) | 64.94 (2.02) | 62.59 (4.14) | p = 0.92 |
| **Age at plasma visit, years** | 65.38 (4.63) | 66.19 (5.18) | 63.70 (0.48) | 64.91 (4.66) | p = 0.79 |
| **Sex, F, n (%)** | 1(13) | 25 (76) | 3 (100) | 36 (73) | p = 0.003 ^a,b,e^ |
| **Education, years** | 13.38 (2.77) | 13.73 (2.55) | 14.67 (4.04) | 15.86 (2.77) | p = 0.006 ^e^ |
| ***APOE* ε4 carriers, n (%)** | 7 (88) | 14 (42) | 2 (67) | 16 (33) | p = 0.02 ^a,c^ |
| **Aβ_42/40_** | 0.08 (0.01) | 0.08 (0.01) | 0.10 (0.02) | 0.10 (0.01) | p < 0.001 ^b,c,d,e^ |
| **pTau217**  **(pg/ml)** | 5.39 (0.98) | 2.15 (0.70) | 5.45 (0.33) | 2.27 (0.67) | p < 0.001 ^a,c,d,f^ |
| **MoCA score**  **/30** | 27.50 (1.41) | 28.06 (1.66) | 29.00 (1.00) | 28.10 (1.34) | p = 0.45 |
| **RBANS**  **global score** | 100.00 (12.27) | 102.42 (10.34) | 95.33 (8.08) | 102.14 (9.11) | p = 0.53 |

**eTable 2. Demographic Characteristics of participants across AT CSF subsample.**

|  | **A+T+** | **A+T-** | **A-T+** | **A-T-** | **Group differences** |
| --- | --- | --- | --- | --- | --- |
|  | **(n = 14)** | **(n = 3)** | **(n = 2)** | **(n = 74)** |  |
| **Age at baseline, years** | 64.41 (5.18) | 68.29 (2.63) | 65.67 (4.80) | 62.69 (4.27) | p = 0.09 |
| **Age at CSF visit, years** | 66.57 (5.58) | 71.37 (1.72) | 69.73 (4.69) | 64.81 (4.50) | p = 0.03 |
| **Sex, F, n (%)** | 6 (43) | 2 (67) | 0 (0) | 57 (77) | p = 0.005 ^c^ |
| **Education, years** | 13.29 (2.89) | 14.00 (3.61) | 18.50 (2.12) | 15.08 (2.78) | p = 0.03 |
| ***APOE* ε4 carriers, n (%)** | 13 (93) | 0 (0) | 1 (50) | 25 (34) | p < 0.001 ^a,c^ |
| **Aβ_42/40_** | 0.05 (0.01) | 0.05 (0.02) | 0.09 (0.01) | 0.10 (0.01) | p < 0.001 ^c,e^ |
| **pTau217**  **(pg/ml)** | 653.29 (119.29) | 310.13 (33.25) | 434.00 (8.59) | 188.92 (79.49) | p < 0.001 ^c^ |
| **MoCA score**  **/30** | 28.00 (1.36) | 28.33 (1.53) | 29.50 (0.71) | 28.03 (1.49) | p = 0.47 |
| **RBANS global score** | 97.07 (11.17) | 105.67 (6.66) | 100.00 (2.83) | 102.64 (9.52) | p = 0.19 |

**eTable 3. Demographic Characteristics of participants across AT PET subsample.**

|  | **A+T+**  **(n = 3)** | **A+T-**  (n = 28) | **A-T+**  **(n = 1)** | A-T-  (n = 61) | **Group differences** |
| --- | --- | --- | --- | --- | --- |
| **Age at baseline, years** | 64.34 (4.64) | 63.63 (4.41) | 62.90 | 62.94 (4.59) | p = 0.72 |
| **Age at PET, years** | 69.53 (4.09) | 67.38 (4.83) | 64.42 | 66.71 (4.62) | p = 0.47 |
| **Sex, F, n (%)** | 2 (67) | 18 (64) | 1(100) | 44 (72) | p = 0.76 |
| **Education, years** | 14.67 (3.79) | 13.82 (2.25) | 11.00 | 15.39 (3.01) | p = 0.05^e^ |
| ***APOE* ε4 carriers, n (%)** | 3 (100) | 16 (57) | 1(100) | 19 (31) | p = 0.007 ^c,e^ |
| **Global Aβ SUVR** | 2.14 (0.30) | 1.51 (0.27) | 1.25 | 1.16 (0.06) | p < 0.001 ^c,e^ |
| **Temporal meta-ROI SUVR** | 1.52 (0.09) | 1.17 (0.07) | 1.55 | 1.13 (0.07) | p < 0.001^a,c,e^ |
| **MoCA score**  **/30** | 28.67 (1.53) | 28.14 (1.60) | 28.00 | 28.00 (1.41) | p = 0.62 |
| **RBANS global score** | 93.00 (12.12) | 101.43 (10.21) | 86.00 | 102.93 (9.22) | p = 0.23 |

Data presented as mean (standard deviation), except for categorical variables where the count and percentage are presented. Fisher or Wallis test were performed between the A/T groups from the subsample (n = 93) and p values are reported on the right column. If a significant group difference (p<0.05) was found post-hoc p values are reported (^a^ difference between A+T+ and A+T-; ^b^ difference between A+T+ and A-T+; ^c^ difference between A+T+ and A-T-; ^d^ difference between A+T- and A-T+; ^e^ A+T- and A-T-; ^f^ A-T+ and A-T-). Age at baseline and plasma collection visit are presented; RBANS values are shown at baseline. MoCA scores were collected at entry into the program. APOE score was missing for one participant in the A+T- group. MoCA score was missing for one participant and RBANS score was missing for four participants in the A-T- group. Abbreviations: MoCA = Montreal Cognitive Assessment; F = female; APOE = apolipoprotein E; SUVR = Standardized Uptake Value Ratio; RBANS = Repeated Battery for for the Assessment of Neuropsychological Status

**eTable 4. Sample Demographics across plasma, CSF and PET full sample.**

| **Demographics** | **Plasma**  **(n = 215)** | **CSF**  **(n = 159)** | **PET**  **(n = 155)** | **Group differences** |
| --- | --- | --- | --- | --- |
| **Age at baseline, years** | 63.19 (4.85) | 62.92 (4.80) | 63.70 (4.62) | p = 0.14 |
| **Age at biomarker classification, years ^a^** | 65.16 (5.28) | 64.75 (5.26) | 67.64 (5.01) | p <0.001* |
| **Sex, F (%)** | 157 (73) | 113 (71) | 111 (72) | p = 0.87 |
| **Education, years** | 15.29 (3.25) | 15.14 (3.17) | 15.33 (3.25) | p = 0.78 |
| ***APOE* ε4 carriers, n (%)** | 88 (41) | 62 (39) | 62 (40) | p = 0.96 |
| **Global amyloid SUVR** | NA | NA | 1.31(0.30) | NA |
| **Temporal meta-ROI SUVR** | NA | NA | 1.16 (0.11) | NA |
| **Aβ_42/40_** | 0.09 (0.01) | 0.09 (0.02) | NA | NA |
| **p-tau217**  **(pg/ml)** | 2.63 (1.56) | 251.70 (173.65) | NA | NA |
| **MoCA score**  **/30 ^b^** | 28.11 (1.57) | 28.05 (1.58) | 28.14 (1.51) | 0.89 |
| **RBANS global score** | 101.61 (9.82) | 101.10 (9.80) | 102.54 (10.12) | 0.22 |

**eTable 5. Demographic Characteristics of participants across AT plasma full sample.**

|  | **A+T+** | **A+T-** | **A-T+** | **A-T-** | **Group differences** |
| --- | --- | --- | --- | --- | --- |
|  | **(n = 21)** | **(n = 60)** | **(n = 7)** | **(n = 127)** |  |
| **Age at baseline, years** | 65.99 (4.90) | 63.99 (5.44) | 64.94 (4.72) | 62.59 (4.41) | p = 0.03^c^ |
| **Age at plasma visit, years** | 68.07 (4.63) | 65.46 (5.83) | 66.53 (5.21) | 64.46 (4.97) | p = 0.01^c^ |
| **Sex, F, n (%)** | 12 (57) | 41 (68) | 6 (86) | 98 (77) | p = 0.17 |
| **Education, years** | 15.10 (3.40) | 14.52 (2.92) | 16 (3.31) | 15.65 (3.31) | p = 0.19 ^c^ |
| ***APOE* ε4 carriers, n (%)** | 16 (76) | 25 (42) | 5 (71) | 42 (33) | p <0.001 ^a,c^ |
| **Aβ_42/40_** | 0.08 (0.01) | 0.08 (0.01) | 0.10 (0.01) | 0.10 (0.01) | p <0.001 ^b,c,d,f^ |
| **pTau217**  **(pg/ml)** | 5.19 (1.05) | 2.19 (0.73) | 6.50 (4.42) | 2.20 (0.74) | p <0.001 ^a,c,d,f^ |
| **MoCA score**  **/30** | 27.67 (1.68) | 28.03 (1.59) | 28.86 (0.90) | 28.17 (1.56) | p = 0.34 |
| **RBANS global score** | 99.41 (12.16) | 101.67 (9.93) | 95.86 (6.59) | 102.19 (9.49) | p = 0.24 |

**eTable 6. Demographic Characteristics of participants across AT CSF full sample.**

|  | **A+T+**  **(n = 18)** | **A+T-**  **(n = 5)** | **A-T+**  **(n = 3)** | **A-T-**  **(n = 133)** | **Group differences** |
| --- | --- | --- | --- | --- | --- |
| **Age at baseline, years** | 63.83 (4.80) | 65.20 (4.98) | 64.54 (3.98) | 62.67 (4.82) | p = 0.33 |
| **Age at CSF visit, years** | 66.30 (5.09) | 67.87(4.97) | 67.92(4.57) | 64.35(5.25) | p = 0.07 |
| **Sex, F, n (%)** | 9 (50) | 3 (60) | 1 (33) | 100 (75) | p = 0.04^c^ |
| **Education, years** | 13.50(2.94) | 14.40 (3.36) | 17.33 (2.52) | 15.35 (3.16) | p = 0.03 |
| ***APOE* ε4 carriers, n (%)** | 16 (89) | 1 (20) | 1 (33) | 44 (44) | p < 0.001 ^a,c^ |
| **Aβ_42/40_** | 0.05 (0.01) | 0.06 (0.01) | 0.09 (0.01) | 0.10 (0.01) | p < 0.001 ^a,c^ |
| **pTau217**  **(pg/ml)** | 663.89(125.89) | 278.88 (48.95) | 426.13 (14.92) | 190.96 (79.82) | p < 0.001 ^c,f^ |
| **MoCA score**  **/30** | 28.17 (1.29) | 28.41 (1.14) | 29.67 (0.58) | 27.98 (1.63) | p = 0.23 |
| **RBANS global score** | 97.94 (10.71) | 105.80 (5.17) | 98.00 (4.00) | 101.42 (9.83) | p = 0.19 |

**eTable 7. Demographic Characteristics of participants across AT PET full sample.**

|  | **A+T+**  **(n = 8)** | **A+T-**  **(n = 45)** | **A-T+**  **(n = 1)** | **A-T-**  **(n = 101)** | **Group differences** |
| --- | --- | --- | --- | --- | --- |
| **Age at baseline, years** | 69.02 (5.24) | 63.66 (4.83) | 62.90 | 63.30 (4.27) | p = 0.02 ^a,c^ |
| **Age at PET, years** | 72.58 (4.22) | 67.41 (5.24) | 64.42 | 67.38 (4.81) | p = 0.02 ^a,c^ |
| **Sex, F, n (%)** | 7 (88) | 33 (73) | 1 (100) | 70 (70) | p = 0.58 |
| **Education, years** | 15.25 (2.60) | 14.62 (2.79) | 11 | 15.69 (3.43) | p = 0.13 |
| ***APOE* ε4 carriers, n (%)** | 6 (75) | 27 (60) | 1(100) | 28 (28) | p < 0.001^c,e^ |
| **Global Aβ SUVR** | 1.98 (0.40) | 1.54 (0.30) | 1.25 | 1.16 (0.06) | p < 0.001^c,e^ |
| **Temporal meta-ROI SUVR** | 1.48 (0.13) | 1.16 (0.06) | 1.55 | 1.12 (0.07) | p < 0.001^a,c,e^ |
| **MoCA score**  **/30** | 28.62 (1.41) | 28.20 (1.66) | 28 | 28.08 (1.47) | p = 0.54 |
| **RBANS global score** | 94.45 (14.65) | 103.33 (9.78) | 86.00 | 102.99 (9.60) | p = 0.34 |

**eTable (4 – 7)** represents the characteristics of plasma, CSF, and PET subsample and their corresponding AT groups. Data presented as mean (standard deviation), except for categorical variables where the count and percentage are presented. Fisher or Wallis test were performed between the groups and p values are reported on the right column. If a significant group difference *(p<0.05) was found post-hoc p values are reported (^a^ difference between A+T+ and A+T-; ^b^ difference between A+T+ and A-T+; ^c^ difference between A+T+ and A-T-; ^d^ difference between A+T- and A-T+; ^e^ A+T- and A-T-; ^f^ A-T+ and A-T-). Age at baseline and PET scan visit are presented; RBANS values are shown at baseline. MoCA scores were collected at entry into the program. APOE was missing for one participant in the A+T- group. MoCA score was missing for one participant and RBANS score was missing for one participant in the A-T- group. Abbreviations: MoCA = Montreal Cognitive Assessment; F = female; APOE = apolipoprotein E; SUVR = Standardized Uptake Value Ratio; RBANS = Repeated Battery for for the Assessment of Neuropsychological Status.

**eTable 8. Diagnostic accuracy of plasma p-tau217 in predicting CI**

**Plasma p-tau217**

**Cutoff	Sensitivity,%	Specificity,%. NPV,%	PPV,%**

| 1.015 | 100 | 1.56 | 100 | 31.52 |
| --- | --- | --- | --- | --- |
| 1.075 | 100 | 3.12 | 100 | 31.87 |
| 1.15 | 100 | 4.69 | 100 | 32.22 |
| 1.235 | 100 | 6.25 | 100 | 32.58 |
| 1.265 | 100 | 7.81 | 100 | 32.95 |
| 1.355 | 96.55 | 7.81 | 83.33 | 32.18 |
| 1.44 | 96.55 | 9.38 | 85.71 | 32.56 |
| 1.47 | 96.55 | 10.94 | 87.5 | 32.94 |
| 1.495 | 96.55 | 12.5 | 88.89 | 33.33 |
| 1.505 | 96.55 | 14.06 | 90 | 33.73 |
| 1.555 | 93.1 | 14.06 | 81.82 | 32.93 |
| 1.625 | 93.1 | 15.62 | 83.33 | 33.33 |
| 1.66 | 93.1 | 17.19 | 84.62 | 33.75 |
| 1.685 | 89.66 | 17.19 | 78.57 | 32.91 |
| 1.705 | 89.66 | 18.75 | 80 | 33.33 |
| 1.72 | 89.66 | 21.88 | 82.35 | 34.21 |
| 1.735 | 89.66 | 23.44 | 83.33 | 34.67 |
| 1.745 | 89.66 | 25 | 84.21 | 35.14 |
| 1.76 | 89.66 | 26.56 | 85 | 35.62 |
| 1.78 | 86.21 | 28.12 | 81.82 | 35.21 |
| 1.8 | 86.21 | 29.69 | 82.61 | 35.71 |
| 1.815 | 86.21 | 32.81 | 84 | 36.76 |
| 1.825 | 86.21 | 34.38 | 84.62 | 37.31 |
| 1.845 | 82.76 | 35.94 | 82.14 | 36.92 |
| 1.87 | 82.76 | 39.06 | 83.33 | 38.1 |
| 1.895 | 82.76 | 40.62 | 83.87 | 38.71 |
| 1.92 | 79.31 | 45.31 | 82.86 | 39.66 |
| 1.94 | 79.31 | 46.88 | 83.33 | 40.35 |
| 1.96 | 75.86 | 46.88 | 81.08 | 39.29 |
| 1.98 | 75.86 | 48.44 | 81.58 | 40 |
| 1.995 | 75.86 | 50 | 82.05 | 40.74 |
| 2.005 | 75.86 | 51.56 | 82.5 | 41.51 |
| 2.015 | 75.86 | 53.12 | 82.93 | 42.31 |
| 2.065 | 72.41 | 53.12 | 80.95 | 41.18 |
| 2.13 | 68.97 | 53.12 | 79.07 | 40 |
| 2.18 | 68.97 | 54.69 | 79.55 | 40.82 |
| 2.235 | 68.97 | 57.81 | 80.43 | 42.55 |
| 2.265 | 68.97 | 59.38 | 80.85 | 43.48 |
| 2.28 | 68.97 | 62.5 | 81.63 | 45.45 |
| 2.295 | 68.97 | 64.06 | 82 | 46.51 |
| 2.315 | 68.97 | 65.62 | 82.35 | 47.62 |
| 2.335 | 65.52 | 65.62 | 80.77 | 46.34 |
| 2.345 | 65.52 | 67.19 | 81.13 | 47.5 |
| 2.365 | 65.52 | 68.75 | 81.48 | 48.72 |
| 2.395 | 65.52 | 70.31 | 81.82 | 50 |
| 2.415 | 65.52 | 71.88 | 82.14 | 51.35 |
| 2.445 | 65.52 | 73.44 | 82.46 | 52.78 |
| 2.48 | 65.52 | 75 | 82.76 | 54.29 |
| 2.505 | 65.52 | 76.56 | 83.05 | 55.88 |
| 2.56 | 65.52 | 78.12 | 83.33 | 57.58 |
| 2.605 | 65.52 | 79.69 | 83.61 | 59.38 |
| 2.635 | 65.52 | 81.25 | 83.87 | 61.29 |
| 2.685 | 65.52 | 82.81 | 84.13 | 63.33 |
| 2.715 | 62.07 | 82.81 | 82.81 | 62.07 |
| 2.735 | 58.62 | 82.81 | 81.54 | 60.71 |
| 2.775 | 51.72 | 82.81 | 79.1 | 57.69 |
| 2.845 | 51.72 | 84.38 | 79.41 | 60 |
| 2.97 | 51.72 | 85.94 | 79.71 | 62.5 |
| 3.105 | 51.72 | 87.5 | 80 | 65.22 |
| 3.175 | 51.72 | 89.06 | 80.28 | 68.18 |
| 3.21 | 51.72 | 90.62 | 80.56 | 71.43 |
| 3.265 | 48.28 | 90.62 | 79.45 | 70 |
| 3.32 | 48.28 | 92.19 | 79.73 | 73.68 |
| 3.38 | 48.28 | 93.75 | 80 | 77.78 |
| 3.485 | 44.83 | 93.75 | 78.95 | 76.47 |
| 3.565 | 44.83 | 95.31 | 79.22 | 81.25 |
| 3.595 | 44.83 | 96.88 | 79.49 | 86.67 |
| 3.615 | 41.38 | 96.88 | 78.48 | 85.71 |
| 3.7 | 37.93 | 96.88 | 77.5 | 84.62 |
| **3.81** | **34.48** | **96.88** | **76.54** | **83.33** |
| 4.27 | 34.48 | 98.44 | 76.83 | 90.91 |
| 4.745 | 31.03 | 100 | 76.19 | 100 |
| 4.815 | 24.14 | 100 | 74.42 | 100 |
| 5.03 | 20.69 | 100 | 73.56 | 100 |
| 5.26 | 17.24 | 100 | 72.73 | 100 |
| 5.38 | 13.79 | 100 | 71.91 | 100 |
| 5.645 | 10.34 | 100 | 71.11 | 100 |
| 6.285 | 6.9 | 100 | 70.33 | 100 |
| 6.915 | 3.45 | 100 | 69.57 | 100 |

**eTable 9. Diagnostic accuracy of plasma Aβ_42/40_ in predicting CI**

**Plasma Aβ_42/40_**

**Cutoff	Sensitivity,% Specificity,%	NPV,%	PPV,%**

| 0.125 | 100 | 1.56 | 100 | 31.52 |
| --- | --- | --- | --- | --- |
| 0.1235 | 96.55 | 1.56 | 50 | 30.77 |
| 0.1225 | 93.1 | 1.56 | 33.33 | 30 |
| 0.121 | 93.1 | 3.12 | 50 | 30.34 |
| 0.1195 | 93.1 | 4.69 | 60 | 30.68 |
| 0.1175 | 93.1 | 6.25 | 66.67 | 31.03 |
| 0.115 | 93.1 | 7.81 | 71.43 | 31.4 |
| 0.113 | 93.1 | 9.38 | 75 | 31.76 |
| 0.111 | 89.66 | 9.38 | 66.67 | 30.95 |
| 0.1095 | 89.66 | 10.94 | 70 | 31.33 |
| 0.1085 | 89.66 | 12.5 | 72.73 | 31.71 |
| 0.1075 | 89.66 | 15.62 | 76.92 | 32.5 |
| 0.1065 | 89.66 | 17.19 | 78.57 | 32.91 |
| 0.1055 | 89.66 | 20.31 | 81.25 | 33.77 |
| 0.1045 | 86.21 | 23.44 | 78.95 | 33.78 |
| 0.103 | 79.31 | 28.12 | 75 | 33.33 |
| 0.1015 | 79.31 | 32.81 | 77.78 | 34.85 |
| 0.1 | 75.86 | 35.94 | 76.67 | 34.92 |
| 0.0985 | 75.86 | 40.62 | 78.79 | 36.67 |
| 0.0975 | 75.86 | 45.31 | 80.56 | 38.6 |
| 0.0965 | 75.86 | 53.12 | 82.93 | 42.31 |
| 0.0955 | 75.86 | 54.69 | 83.33 | 43.14 |
| 0.0945 | 75.86 | 57.81 | 84.09 | 44.9 |
| 0.0935 | 72.41 | 59.38 | 82.61 | 44.68 |
| 0.0925 | 72.41 | 64.06 | 83.67 | 47.73 |
| 0.0915 | 68.97 | 65.62 | 82.35 | 47.62 |
| **0.0905** | **65.52** | **65.62** | **80.77** | **46.34** |
| 0.0895 | 65.52 | 68.75 | 81.48 | 48.72 |
| 0.0885 | 62.07 | 70.31 | 80.36 | 48.65 |
| 0.087 | 55.17 | 71.88 | 77.97 | 47.06 |
| 0.0855 | 51.72 | 75 | 77.42 | 48.39 |
| 0.0845 | 48.28 | 78.12 | 76.92 | 50 |
| 0.0835 | 37.93 | 81.25 | 74.29 | 47.83 |
| 0.082 | 34.48 | 84.38 | 73.97 | 50 |
| 0.0805 | 27.59 | 85.94 | 72.37 | 47.06 |
| 0.0795 | 24.14 | 85.94 | 71.43 | 43.75 |
| 0.0785 | 24.14 | 87.5 | 71.79 | 46.67 |
| 0.0775 | 20.69 | 89.06 | 71.25 | 46.15 |
| 0.0765 | 20.69 | 90.62 | 71.6 | 50 |
| 0.0755 | 20.69 | 92.19 | 71.95 | 54.55 |
| 0.074 | 17.24 | 95.31 | 71.76 | 62.5 |
| 0.0725 | 13.79 | 95.31 | 70.93 | 57.14 |
| 0.0715 | 10.34 | 95.31 | 70.11 | 50 |
| 0.0695 | 3.45 | 96.88 | 68.89 | 33.33 |
| 0.067 | 0 | 96.88 | 68.13 | 0 |
| 0.0485 | 0 | 98.44 | 68.48 | 0 |

**eTable 10. Diagnostic accuracy of CSF p-tau217 in predicting CI**

| **CSF p-tau217** | | | | |
| --- | --- | --- | --- | --- |
| Cutoff | Sensitivity,% | Specificity,% | NPV,% | PPV,% |
| 76.905 | 100 | 1.56 | 100 | 31.52 |
| 82.545 | 100 | 3.12 | 100 | 31.87 |
| 84.945 | 100 | 4.69 | 100 | 32.22 |
| 86.87 | 100 | 6.25 | 100 | 32.58 |
| 87.895 | 100 | 7.81 | 100 | 32.95 |
| 89.52 | 100 | 9.38 | 100 | 33.33 |
| 91.465 | 100 | 10.94 | 100 | 33.72 |
| 92.845 | 100 | 12.5 | 100 | 34.12 |
| 95.275 | 100 | 14.06 | 100 | 34.52 |
| 98.935 | 96.55 | 14.06 | 90 | 33.73 |
| 101.08 | 96.55 | 15.62 | 90.91 | 34.15 |
| 101.48 | 96.55 | 17.19 | 91.67 | 34.57 |
| 102.105 | 96.55 | 18.75 | 92.31 | 35 |
| 106.965 | 96.55 | 20.31 | 92.86 | 35.44 |
| 112.345 | 93.1 | 20.31 | 86.67 | 34.62 |
| 114.815 | 93.1 | 21.88 | 87.5 | 35.06 |
| 120.135 | 93.1 | 23.44 | 88.24 | 35.53 |
| 125.145 | 93.1 | 25 | 88.89 | 36 |
| 127.915 | 93.1 | 26.56 | 89.47 | 36.49 |
| 131.105 | 93.1 | 28.12 | 90 | 36.99 |
| 133.045 | 93.1 | 29.69 | 90.48 | 37.5 |
| 133.65 | 93.1 | 31.25 | 90.91 | 38.03 |
| 135.37 | 89.66 | 31.25 | 86.96 | 37.14 |
| 139.7 | 89.66 | 32.81 | 87.5 | 37.68 |
| 145.89 | 89.66 | 34.38 | 88 | 38.24 |
| 149.18 | 89.66 | 35.94 | 88.46 | 38.81 |
| 151.57 | 89.66 | 37.5 | 88.89 | 39.39 |
| 154.39 | 89.66 | 39.06 | 89.29 | 40 |
| 155.355 | 89.66 | 40.62 | 89.66 | 40.62 |
| 156.13 | 89.66 | 42.19 | 90 | 41.27 |
| 158.265 | 86.21 | 42.19 | 87.1 | 40.32 |
| 161.155 | 86.21 | 45.31 | 87.88 | 41.67 |
| 163.33 | 86.21 | 46.88 | 88.24 | 42.37 |
| 166.215 | 86.21 | 48.44 | 88.57 | 43.1 |
| 170.11 | 86.21 | 50 | 88.89 | 43.86 |
| 173.11 | 86.21 | 51.56 | 89.19 | 44.64 |
| 176.05 | 86.21 | 53.12 | 89.47 | 45.45 |
| 180.57 | 86.21 | 54.69 | 89.74 | 46.3 |
| 184.54 | 86.21 | 56.25 | 90 | 47.17 |
| 186.625 | 82.76 | 56.25 | 87.8 | 46.15 |
| 188.45 | 82.76 | 57.81 | 88.1 | 47.06 |
| 196.42 | 82.76 | 59.38 | 88.37 | 48 |
| 206.71 | 79.31 | 59.38 | 86.36 | 46.94 |
| 213.51 | 79.31 | 60.94 | 86.67 | 47.92 |
| 217.335 | 79.31 | 62.5 | 86.96 | 48.94 |
| 218.865 | 79.31 | 64.06 | 87.23 | 50 |
| 219.99 | 79.31 | 65.62 | 87.5 | 51.11 |
| 220.355 | 79.31 | 67.19 | 87.76 | 52.27 |
| 223.14 | 75.86 | 67.19 | 86 | 51.16 |
| 225.735 | 75.86 | 68.75 | 86.27 | 52.38 |
| 226.4 | 75.86 | 70.31 | 86.54 | 53.66 |
| 228.635 | 75.86 | 71.88 | 86.79 | 55 |
| 230.28 | 72.41 | 71.88 | 85.19 | 53.85 |
| 234.345 | 72.41 | 73.44 | 85.45 | 55.26 |
| 238.52 | 72.41 | 75 | 85.71 | 56.76 |
| 240.045 | 72.41 | 76.56 | 85.96 | 58.33 |
| 243.25 | 72.41 | 78.12 | 86.21 | 60 |
| 249.11 | 72.41 | 79.69 | 86.44 | 61.76 |
| 256.365 | 72.41 | 81.25 | 86.67 | 63.64 |
| 261.555 | 72.41 | 82.81 | 86.89 | 65.62 |
| 272.12 | 68.97 | 82.81 | 85.48 | 64.52 |
| 281.975 | 65.52 | 82.81 | 84.13 | 63.33 |
| 285.095 | 65.52 | 84.38 | 84.38 | 65.52 |
| 287.205 | 65.52 | 85.94 | 84.62 | 67.86 |
| 293.655 | 62.07 | 85.94 | 83.33 | 66.67 |
| 306.445 | 58.62 | 85.94 | 82.09 | 65.38 |
| 316.54 | 55.17 | 85.94 | 80.88 | 64 |
| 320.625 | 55.17 | 87.5 | 81.16 | 66.67 |
| 323.37 | 55.17 | 89.06 | 81.43 | 69.57 |
| 335.92 | 55.17 | 90.62 | 81.69 | 72.73 |
| 346.785 | 51.72 | 90.62 | 80.56 | 71.43 |
| 347.54 | 51.72 | 92.19 | 80.82 | 75 |
| 351.04 | 48.28 | 92.19 | 79.73 | 73.68 |
| 357.425 | 48.28 | 93.75 | 80 | 77.78 |
| 362.57 | 44.83 | 93.75 | 78.95 | 76.47 |
| **396.255** | **44.83** | **95.31** | **79.22** | **81.25** |
| 433.995 | 44.83 | 96.88 | 79.49 | 86.67 |
| 498.345 | 41.38 | 96.88 | 78.48 | 85.71 |
| 560.94 | 41.38 | 98.44 | 78.75 | 92.31 |
| 569.905 | 37.93 | 98.44 | 77.78 | 91.67 |
| 579.18 | 34.48 | 98.44 | 76.83 | 90.91 |
| 589.27 | 31.03 | 98.44 | 75.9 | 90 |
| 597.845 | 27.59 | 98.44 | 75 | 88.89 |
| 605.82 | 24.14 | 98.44 | 74.12 | 87.5 |
| 611.95 | 20.69 | 98.44 | 73.26 | 85.71 |
| 626.95 | 17.24 | 98.44 | 72.41 | 83.33 |
| 647.61 | 13.79 | 98.44 | 71.59 | 80 |
| 672.005 | 10.34 | 98.44 | 70.79 | 75 |
| 693.45 | 6.9 | 98.44 | 70 | 66.67 |
| 719.715 | 3.45 | 98.44 | 69.23 | 50 |
| 882.04 | 3.45 | 100 | 69.57 | 100 |

**eTable 11. Diagnostic accuracy of CSF Aβ_42/40_ in predicting CI**

**CSF Aβ_42/40_**

**Cutoff	Sensitivity,%	Specificity,%	NPV,%	PPV,%**

| 0.11755925 | 100 | 1.56 | 100 | 31.52 |
| --- | --- | --- | --- | --- |
| 0.11629457 | 100 | 3.12 | 100 | 31.87 |
| 0.11550666 | 100 | 4.69 | 100 | 32.22 |
| 0.11520654 | 100 | 6.25 | 100 | 32.58 |
| 0.11467891 | 96.55 | 6.25 | 80 | 31.82 |
| 0.11431674 | 93.1 | 6.25 | 66.67 | 31.03 |
| 0.11387694 | 93.1 | 7.81 | 71.43 | 31.4 |
| 0.11314396 | 93.1 | 9.38 | 75 | 31.76 |
| 0.11241106 | 93.1 | 10.94 | 77.78 | 32.14 |
| 0.1115638 | 93.1 | 12.5 | 80 | 32.53 |
| 0.11078507 | 89.66 | 12.5 | 72.73 | 31.71 |
| 0.11041892 | 89.66 | 14.06 | 75 | 32.1 |
| 0.1103296 | 89.66 | 15.62 | 76.92 | 32.5 |
| 0.11002579 | 89.66 | 17.19 | 78.57 | 32.91 |
| 0.1097942 | 89.66 | 18.75 | 80 | 33.33 |
| 0.1094816 | 89.66 | 20.31 | 81.25 | 33.77 |
| 0.10891789 | 89.66 | 21.88 | 82.35 | 34.21 |
| 0.10865698 | 86.21 | 21.88 | 77.78 | 33.33 |
| 0.10836479 | 86.21 | 23.44 | 78.95 | 33.78 |
| 0.10793686 | 86.21 | 25 | 80 | 34.25 |
| 0.10771086 | 82.76 | 25 | 76.19 | 33.33 |
| 0.10761779 | 82.76 | 26.56 | 77.27 | 33.8 |
| 0.10757785 | 79.31 | 26.56 | 73.91 | 32.86 |
| 0.10733048 | 75.86 | 26.56 | 70.83 | 31.88 |
| 0.1071193 | 75.86 | 28.12 | 72 | 32.35 |
| 0.10698218 | 75.86 | 29.69 | 73.08 | 32.84 |
| 0.106841 | 75.86 | 31.25 | 74.07 | 33.33 |
| 0.10667824 | 75.86 | 32.81 | 75 | 33.85 |
| 0.10597734 | 75.86 | 34.38 | 75.86 | 34.38 |
| 0.10542111 | 75.86 | 35.94 | 76.67 | 34.92 |
| 0.10515465 | 75.86 | 37.5 | 77.42 | 35.48 |
| 0.10487495 | 75.86 | 39.06 | 78.12 | 36.07 |
| 0.10473536 | 72.41 | 39.06 | 75.76 | 35 |
| 0.10460357 | 72.41 | 40.62 | 76.47 | 35.59 |
| 0.10387171 | 68.97 | 40.62 | 74.29 | 34.48 |
| 0.10312531 | 68.97 | 42.19 | 75 | 35.09 |
| 0.10245297 | 68.97 | 43.75 | 75.68 | 35.71 |
| 0.10177143 | 68.97 | 45.31 | 76.32 | 36.36 |
| 0.10163765 | 68.97 | 46.88 | 76.92 | 37.04 |
| 0.10123674 | 68.97 | 48.44 | 77.5 | 37.74 |
| 0.10078462 | 68.97 | 50 | 78.05 | 38.46 |
| 0.10052623 | 68.97 | 51.56 | 78.57 | 39.22 |
| 0.09991946 | 68.97 | 53.12 | 79.07 | 40 |
| 0.0993973 | 68.97 | 54.69 | 79.55 | 40.82 |
| 0.09935234 | 65.52 | 54.69 | 77.78 | 39.58 |
| 0.09921656 | 65.52 | 56.25 | 78.26 | 40.43 |
| 0.09807373 | 62.07 | 56.25 | 76.6 | 39.13 |
| 0.09658938 | 62.07 | 57.81 | 77.08 | 40 |
| 0.09594203 | 62.07 | 59.38 | 77.55 | 40.91 |
| 0.09572916 | 58.62 | 59.38 | 76 | 39.53 |
| 0.09561533 | 58.62 | 60.94 | 76.47 | 40.48 |
| 0.09544816 | 58.62 | 62.5 | 76.92 | 41.46 |
| 0.09527615 | 58.62 | 64.06 | 77.36 | 42.5 |
| 0.09475347 | 58.62 | 65.62 | 77.78 | 43.59 |
| 0.09430784 | 58.62 | 67.19 | 78.18 | 44.74 |
| 0.0941041 | 58.62 | 68.75 | 78.57 | 45.95 |
| 0.09372267 | 58.62 | 70.31 | 78.95 | 47.22 |
| 0.09298958 | 58.62 | 71.88 | 79.31 | 48.57 |
| 0.09123886 | 58.62 | 73.44 | 79.66 | 50 |
| 0.08992333 | 58.62 | 75 | 80 | 51.52 |
| 0.0897631 | 58.62 | 76.56 | 80.33 | 53.12 |
| 0.08952656 | 58.62 | 78.12 | 80.65 | 54.84 |
| 0.08891739 | 58.62 | 79.69 | 80.95 | 56.67 |
| 0.08831979 | 55.17 | 79.69 | 79.69 | 55.17 |
| 0.08789368 | 51.72 | 79.69 | 78.46 | 53.57 |
| 0.08757762 | 51.72 | 81.25 | 78.79 | 55.56 |
| 0.08747697 | 48.28 | 81.25 | 77.61 | 53.85 |
| 0.08688775 | 48.28 | 82.81 | 77.94 | 56 |
| 0.08346913 | 48.28 | 84.38 | 78.26 | 58.33 |
| 0.07954653 | 48.28 | 85.94 | 78.57 | 60.87 |
| 0.07816239 | 48.28 | 87.5 | 78.87 | 63.64 |
| 0.07781973 | 44.83 | 87.5 | 77.78 | 61.9 |
| 0.07774518 | 44.83 | 89.06 | 78.08 | 65 |
| 0.0770715 | 44.83 | 90.62 | 78.38 | 68.42 |
| 0.07589199 | 44.83 | 92.19 | 78.67 | 72.22 |
| **0.07027368** | **44.83** | **93.75** | **78.95** | **76.47** |
| 0.06507624 | 44.83 | 95.31 | 79.22 | 81.25 |
| 0.06481599 | 41.38 | 95.31 | 78.21 | 80 |
| 0.06305217 | 41.38 | 96.88 | 78.48 | 85.71 |
| 0.05935628 | 37.93 | 96.88 | 77.5 | 84.62 |
| 0.05722767 | 34.48 | 96.88 | 76.54 | 83.33 |
| 0.05699247 | 31.03 | 96.88 | 75.61 | 81.82 |
| 0.05495576 | 27.59 | 96.88 | 74.7 | 80 |
| 0.05300843 | 24.14 | 96.88 | 73.81 | 77.78 |
| 0.0522687 | 20.69 | 96.88 | 72.94 | 75 |
| 0.04782442 | 17.24 | 96.88 | 72.09 | 71.43 |
| 0.04401495 | 13.79 | 96.88 | 71.26 | 66.67 |
| 0.04304896 | 10.34 | 96.88 | 70.45 | 60 |
| 0.04109527 | 6.9 | 96.88 | 69.66 | 50 |
| 0.03958742 | 3.45 | 96.88 | 68.89 | 33.33 |
| 0.03815797 | 3.45 | 98.44 | 69.23 | 50 |
| 0.03708082 | 0 | 98.44 | 68.48 | 0 |

**eTable 12. Diagnostic accuracy of metaROI tau-PET in predicting CI**

**metaROI tau-PET SUVR**

**Cutoff	Sensitivity,%	Specificity,%. NPV,%	PPV,%**

| 0.93387917 | 100 | 1.56 | 100 | 31.52 |
| --- | --- | --- | --- | --- |
| 1.02888333 | 100 | 3.12 | 100 | 31.87 |
| 1.03296667 | 100 | 4.69 | 100 | 32.22 |
| 1.04064583 | 100 | 6.25 | 100 | 32.58 |
| 1.04680833 | 96.55 | 6.25 | 80 | 31.82 |
| 1.05005833 | 96.55 | 7.81 | 83.33 | 32.18 |
| 1.05174583 | 96.55 | 9.38 | 85.71 | 32.56 |
| 1.05585833 | 96.55 | 10.94 | 87.5 | 32.94 |
| 1.059225 | 96.55 | 12.5 | 88.89 | 33.33 |
| 1.06004167 | 96.55 | 14.06 | 90 | 33.73 |
| 1.06235833 | 96.55 | 15.62 | 90.91 | 34.15 |
| 1.06590417 | 96.55 | 17.19 | 91.67 | 34.57 |
| 1.07028333 | 96.55 | 18.75 | 92.31 | 35 |
| 1.07317083 | 96.55 | 20.31 | 92.86 | 35.44 |
| 1.07369167 | 96.55 | 21.88 | 93.33 | 35.9 |
| 1.07482083 | 96.55 | 23.44 | 93.75 | 36.36 |
| 1.0761625 | 96.55 | 25 | 94.12 | 36.84 |
| 1.07802917 | 96.55 | 26.56 | 94.44 | 37.33 |
| 1.08136667 | 93.1 | 26.56 | 89.47 | 36.49 |
| 1.08465 | 93.1 | 28.12 | 90 | 36.99 |
| 1.0876125 | 93.1 | 29.69 | 90.48 | 37.5 |
| 1.091425 | 93.1 | 31.25 | 90.91 | 38.03 |
| 1.09397083 | 93.1 | 32.81 | 91.3 | 38.57 |
| 1.09449583 | 93.1 | 34.38 | 91.67 | 39.13 |
| 1.0954875 | 89.66 | 34.38 | 88 | 38.24 |
| 1.0968375 | 89.66 | 35.94 | 88.46 | 38.81 |
| 1.09965417 | 86.21 | 35.94 | 85.19 | 37.88 |
| 1.1036625 | 82.76 | 35.94 | 82.14 | 36.92 |
| 1.105975 | 79.31 | 35.94 | 79.31 | 35.94 |
| 1.10824583 | 79.31 | 37.5 | 80 | 36.51 |
| 1.11210833 | 79.31 | 39.06 | 80.65 | 37.1 |
| 1.11415417 | 79.31 | 40.62 | 81.25 | 37.7 |
| 1.11581667 | 75.86 | 40.62 | 78.79 | 36.67 |
| 1.11866667 | 75.86 | 42.19 | 79.41 | 37.29 |
| 1.12019167 | 75.86 | 43.75 | 80 | 37.93 |
| 1.12160417 | 75.86 | 45.31 | 80.56 | 38.6 |
| 1.12307917 | 75.86 | 46.88 | 81.08 | 39.29 |
| 1.1238375 | 75.86 | 48.44 | 81.58 | 40 |
| 1.12503333 | 75.86 | 50 | 82.05 | 40.74 |
| 1.12792917 | 75.86 | 51.56 | 82.5 | 41.51 |
| 1.130375 | 75.86 | 53.12 | 82.93 | 42.31 |
| 1.13249167 | 72.41 | 53.12 | 80.95 | 41.18 |
| 1.13436667 | 72.41 | 54.69 | 81.4 | 42 |
| 1.1346 | 72.41 | 56.25 | 81.82 | 42.86 |
| 1.13725833 | 72.41 | 57.81 | 82.22 | 43.75 |
| 1.14160417 | 72.41 | 59.38 | 82.61 | 44.68 |
| 1.14356667 | 72.41 | 60.94 | 82.98 | 45.65 |
| 1.14482917 | 72.41 | 62.5 | 83.33 | 46.67 |
| 1.14882083 | 68.97 | 62.5 | 81.63 | 45.45 |
| 1.15248333 | 68.97 | 64.06 | 82 | 46.51 |
| 1.15384583 | 68.97 | 65.62 | 82.35 | 47.62 |
| 1.1560875 | 68.97 | 67.19 | 82.69 | 48.78 |
| 1.15800833 | 68.97 | 68.75 | 83.02 | 50 |
| 1.16129167 | 68.97 | 70.31 | 83.33 | 51.28 |
| 1.1653 | 68.97 | 71.88 | 83.64 | 52.63 |
| 1.166475 | 68.97 | 73.44 | 83.93 | 54.05 |
| 1.16797083 | 65.52 | 73.44 | 82.46 | 52.78 |
| 1.16966667 | 65.52 | 75 | 82.76 | 54.29 |
| 1.17177917 | 65.52 | 76.56 | 83.05 | 55.88 |
| 1.17515417 | 65.52 | 78.12 | 83.33 | 57.58 |
| 1.17679167 | 62.07 | 78.12 | 81.97 | 56.25 |
| 1.17692083 | 58.62 | 78.12 | 80.65 | 54.84 |
| 1.178625 | 55.17 | 78.12 | 79.37 | 53.33 |
| 1.18093333 | 51.72 | 78.12 | 78.12 | 51.72 |
| 1.18207917 | 51.72 | 79.69 | 78.46 | 53.57 |
| 1.1826125 | 51.72 | 81.25 | 78.79 | 55.56 |
| 1.18456667 | 51.72 | 82.81 | 79.1 | 57.69 |
| 1.18687083 | 51.72 | 84.38 | 79.41 | 60 |
| 1.18985417 | 48.28 | 84.38 | 78.26 | 58.33 |
| 1.19388333 | 44.83 | 84.38 | 77.14 | 56.52 |
| 1.19658333 | 41.38 | 84.38 | 76.06 | 54.55 |
| 1.2024625 | 41.38 | 85.94 | 76.39 | 57.14 |
| 1.20722917 | 41.38 | 87.5 | 76.71 | 60 |
| 1.20845 | 37.93 | 87.5 | 75.68 | 57.89 |
| 1.214325 | 37.93 | 89.06 | 76 | 61.11 |
| 1.2193875 | 37.93 | 90.62 | 76.32 | 64.71 |
| 1.22206667 | 37.93 | 92.19 | 76.62 | 68.75 |
| 1.2259625 | 34.48 | 92.19 | 75.64 | 66.67 |
| 1.22948333 | 34.48 | 93.75 | 75.95 | 71.43 |
| 1.231575 | 31.03 | 93.75 | 75 | 69.23 |
| 1.23369583 | 27.59 | 93.75 | 74.07 | 66.67 |
| 1.2361 | 27.59 | 95.31 | 74.39 | 72.73 |
| 1.24079167 | 27.59 | 96.88 | 74.7 | 80 |
| 1.24540833 | 27.59 | 98.44 | 75 | 88.89 |
| 1.24978333 | 24.14 | 98.44 | 74.12 | 87.5 |
| 1.25456667 | 20.69 | 98.44 | 73.26 | 85.71 |
| 1.2602375 | 20.69 | 100 | 73.56 | 100 |
| **1.26750833** | **17.24** | **100** | **72.73** | **100** |
| 1.35673333 | 13.79 | 100 | 71.91 | 100 |
| 1.46423333 | 10.34 | 100 | 71.11 | 100 |
| 1.51714583 | 6.9 | 100 | 70.33 | 100 |
| 1.58541667 | 3.45 | 100 | 69.57 | 100 |

**eTable 13. Diagnostic accuracy of Aβ -PET in predicting CI**

**Global Aβ -PET SUVR**

**Cutoff	Sensitivity, %	Specificity,%	NPV,%	PPV,%**

| 1.0291 | 100 | 1.56 | 100 | 31.52 |
| --- | --- | --- | --- | --- |
| 1.0436 | 100 | 3.12 | 100 | 31.87 |
| 1.0659 | 100 | 4.69 | 100 | 32.22 |
| 1.0768 | 100 | 6.25 | 100 | 32.58 |
| 1.07995 | 96.55 | 6.25 | 80 | 31.82 |
| 1.0824 | 96.55 | 7.81 | 83.33 | 32.18 |
| 1.08475 | 96.55 | 9.38 | 85.71 | 32.56 |
| 1.0883 | 96.55 | 10.94 | 87.5 | 32.94 |
| 1.09195 | 96.55 | 12.5 | 88.89 | 33.33 |
| 1.09425 | 96.55 | 14.06 | 90 | 33.73 |
| 1.10765 | 96.55 | 15.62 | 90.91 | 34.15 |
| 1.11945 | 96.55 | 17.19 | 91.67 | 34.57 |
| 1.1206 | 96.55 | 18.75 | 92.31 | 35 |
| 1.12225 | 96.55 | 20.31 | 92.86 | 35.44 |
| 1.1269 | 96.55 | 21.88 | 93.33 | 35.9 |
| 1.13645 | 96.55 | 23.44 | 93.75 | 36.36 |
| 1.1441 | 93.1 | 23.44 | 88.24 | 35.53 |
| 1.1466 | 93.1 | 25 | 88.89 | 36 |
| 1.14845 | 93.1 | 26.56 | 89.47 | 36.49 |
| 1.14965 | 93.1 | 28.12 | 90 | 36.99 |
| 1.15115 | 93.1 | 29.69 | 90.48 | 37.5 |
| 1.1529 | 93.1 | 31.25 | 90.91 | 38.03 |
| 1.15345 | 93.1 | 32.81 | 91.3 | 38.57 |
| 1.1537 | 89.66 | 32.81 | 87.5 | 37.68 |
| 1.1546 | 86.21 | 32.81 | 84 | 36.76 |
| 1.1555 | 86.21 | 34.38 | 84.62 | 37.31 |
| 1.15585 | 86.21 | 35.94 | 85.19 | 37.88 |
| 1.15765 | 86.21 | 37.5 | 85.71 | 38.46 |
| 1.15945 | 86.21 | 39.06 | 86.21 | 39.06 |
| 1.15985 | 86.21 | 40.62 | 86.67 | 39.68 |
| 1.16065 | 86.21 | 42.19 | 87.1 | 40.32 |
| 1.162 | 86.21 | 43.75 | 87.5 | 40.98 |
| 1.16395 | 86.21 | 45.31 | 87.88 | 41.67 |
| 1.16525 | 86.21 | 46.88 | 88.24 | 42.37 |
| 1.1656 | 82.76 | 46.88 | 85.71 | 41.38 |
| 1.16625 | 82.76 | 48.44 | 86.11 | 42.11 |
| 1.17465 | 82.76 | 50 | 86.49 | 42.86 |
| 1.18295 | 82.76 | 51.56 | 86.84 | 43.64 |
| 1.187 | 82.76 | 53.12 | 87.18 | 44.44 |
| 1.19345 | 79.31 | 53.12 | 85 | 43.4 |
| 1.1976 | 79.31 | 54.69 | 85.37 | 44.23 |
| 1.2003 | 75.86 | 54.69 | 83.33 | 43.14 |
| 1.203 | 75.86 | 56.25 | 83.72 | 44 |
| 1.20465 | 75.86 | 57.81 | 84.09 | 44.9 |
| 1.20495 | 75.86 | 59.38 | 84.44 | 45.83 |
| 1.2052 | 72.41 | 59.38 | 82.61 | 44.68 |
| 1.2056 | 68.97 | 59.38 | 80.85 | 43.48 |
| 1.20705 | 65.52 | 59.38 | 79.17 | 42.22 |
| 1.2088 | 65.52 | 60.94 | 79.59 | 43.18 |
| 1.21065 | 62.07 | 60.94 | 78 | 41.86 |
| 1.21325 | 62.07 | 62.5 | 78.43 | 42.86 |
| 1.21585 | 62.07 | 64.06 | 78.85 | 43.9 |
| 1.21805 | 62.07 | 65.62 | 79.25 | 45 |
| 1.22365 | 62.07 | 67.19 | 79.63 | 46.15 |
| 1.23405 | 62.07 | 68.75 | 80 | 47.37 |
| 1.24325 | 62.07 | 70.31 | 80.36 | 48.65 |
| 1.24695 | 62.07 | 71.88 | 80.7 | 50 |
| 1.24755 | 62.07 | 73.44 | 81.03 | 51.43 |
| 1.2482 | 58.62 | 73.44 | 79.66 | 50 |
| 1.2493 | 58.62 | 75 | 80 | 51.52 |
| 1.25435 | 55.17 | 75 | 78.69 | 50 |
| 1.266 | 55.17 | 76.56 | 79.03 | 51.61 |
| **1.27355** | **51.72** | **76.56** | **77.78** | **50** |
| 1.27465 | 51.72 | 78.12 | 78.12 | 51.72 |
| 1.27925 | 51.72 | 79.69 | 78.46 | 53.57 |
| 1.2881 | 51.72 | 81.25 | 78.79 | 55.56 |
| 1.29385 | 48.28 | 81.25 | 77.61 | 53.85 |
| 1.29545 | 48.28 | 82.81 | 77.94 | 56 |
| 1.29725 | 48.28 | 84.38 | 78.26 | 58.33 |
| 1.3088 | 48.28 | 85.94 | 78.57 | 60.87 |
| 1.32835 | 48.28 | 87.5 | 78.87 | 63.64 |
| 1.349 | 44.83 | 87.5 | 77.78 | 61.9 |
| 1.3645 | 44.83 | 89.06 | 78.08 | 65 |
| 1.37455 | 44.83 | 90.62 | 78.38 | 68.42 |
| 1.38505 | 41.38 | 90.62 | 77.33 | 66.67 |
| 1.4041 | 41.38 | 92.19 | 77.63 | 70.59 |
| 1.4214 | 41.38 | 93.75 | 77.92 | 75 |
| 1.4348 | 41.38 | 95.31 | 78.21 | 80 |
| 1.4788 | 41.38 | 96.88 | 78.48 | 85.71 |
| 1.51825 | 37.93 | 96.88 | 77.5 | 84.62 |
| 1.56195 | 37.93 | 98.44 | 77.78 | 91.67 |
| 1.63095 | 34.48 | 98.44 | 76.83 | 90.91 |
| 1.66685 | 31.03 | 98.44 | 75.9 | 90 |
| 1.6735 | 27.59 | 98.44 | 75 | 88.89 |
| 1.70335 | 27.59 | 100 | 75.29 | 100 |
| 1.764 | 24.14 | 100 | 74.42 | 100 |
| 1.8415 | 20.69 | 100 | 73.56 | 100 |
| 1.89045 | 17.24 | 100 | 72.73 | 100 |
| 1.9434 | 13.79 | 100 | 71.91 | 100 |
| 2.11375 | 10.34 | 100 | 71.11 | 100 |
| 2.3088 | 6.9 | 100 | 70.33 | 100 |
| 2.39375 | 3.45 | 100 | 69.57 | 100 |

**eTable 8-13.** Comparison estimates of fluid biomarkers vs PET imaging in predicting CI. The sensitivity, specificity, positive predictive values, and negative predictive values of the cutoffs used in the manuscript are presented in bold. Abbreviations: PPV = positive predictive value, NPV = negative predictive value.

**eReferences**

1.	Tremblay-Mercier J, Madjar C, Das S, et al. Open science datasets from PREVENT-AD, a longitudinal cohort of pre-symptomatic Alzheimer's disease. *NeuroImage Clinical*. 2021;31:102733. doi:10.1016/j.nicl.2021.102733

2.	Meyer P-F, Ashton NJ, Karikari TK, et al. Plasma p-tau231, p-tau181, PET Biomarkers, and Cognitive Change in Older Adults. *Annals of neurology*. 2022;91(4):548-560. doi:10.1002/ana.26308

3.	Gonzalez-Ortiz F, Ferreira PCL, González-Escalante A, et al. A novel ultrasensitive assay for plasma p-tau217: Performance in individuals with subjective cognitive decline and early Alzheimer's disease. *Alzheimers Dement*. Feb 2024;20(2):1239-1249. doi:10.1002/alz.13525

4.	Meyer P-F, Savard M, Poirier J, et al. Bi-directional Association of Cerebrospinal Fluid Immune Markers with Stage of&nbsp;Alzheimer’s Disease Pathogenesis. *Journal of Alzheimer's Disease*. 2018;63:577-590. doi:10.3233/JAD-170887

5.	Gobom J, Parnetti L, Rosa-Neto P, et al. Validation of the LUMIPULSE automated immunoassay for the measurement of core AD biomarkers in cerebrospinal fluid. *Clin Chem Lab Med*. Jan 27 2022;60(2):207-219. doi:10.1515/cclm-2021-0651

6.	Sibomana M, Keller S, Stute S, Comtat C. *Benefits of 3D scatter correction for the HRRT - a large axial FOV PET scanner*. 2012:2954-2957.

7.	Desikan RS, Ségonne F, Fischl B, et al. An automated labeling system for subdividing the human cerebral cortex on MRI scans into gyral based regions of interest. *NeuroImage*. 2006;31(3):968-80.

8.	Villeneuve S, Rabinovici GD, Cohn-Sheehy BI, et al. Existing Pittsburgh Compound-B positron emission tomography thresholds are too high: statistical and pathological evaluation. 2015;138(7):2020-2033. doi:10.1093/brain/awv112

9.	Baker SL, Maass A, Jagust WJ, Lawrence Berkeley National Lab BCA. Considerations and code for partial volume correcting [ 18 F]-AV-1451 tau PET data. *Data in Brief*. 2017;15(C)doi:10.1016/j.dib.2017.10.024

10.	Strikwerda-Brown C, Hobbs DA, Gonneaud J, et al. Association of Elevated Amyloid and Tau Positron Emission Tomography Signal With Near-Term Development of Alzheimer Disease Symptoms in Older Adults Without Cognitive Impairment. *JAMA Neurology*. 2022;doi:10.1001/jamaneurol.2022.2379
